# Effectiveness and durability of protection against future SARS-CoV-2 infection conferred by COVID-19 vaccination and previous infection; findings from the UK SIREN prospective cohort study of healthcare workers March 2020 to September 2021

**DOI:** 10.1101/2021.11.29.21267006

**Authors:** Victoria Hall, Sarah Foulkes, Ferdinando Insalata, Ayoub Saei, Peter Kirwan, Ana Atti, Edgar Wellington, Jameel Khawam, Katie Munro, Michelle Cole, Caio Tranquillini, Andrew Taylor-Kerr, Nipunadi Hettiarachchi, Davina Calbraith, Noshin Sajedi, Iain Milligan, Yrene Themistocleous, Diane Corrigan, Lisa Cromey, Lesley Price, Sally Stewart, Elen de Lacy, Chris Norman, Ezra Linley, Ashley David Otter, Amanda Semper, Jacqueline Hewson, Silvia D’Arcangelo, the SIREN Study Group, Meera Chand, Colin S Brown, Tim Brooks, Jasmin Islam, Andre Charlett, Susan Hopkins

## Abstract

**Background:** Understanding the duration and effectiveness of infection and vaccine-acquired SARS-CoV-2 immunity is essential to inform pandemic policy interventions, including the timing of vaccine-boosters. We investigated this in our large prospective cohort of UK healthcare workers undergoing routine asymptomatic PCR testing.

**Methods:** We assessed vaccine effectiveness (VE) (up to 10-months after first dose) and infection-acquired immunity by comparing time to PCR-confirmed infection in vaccinated and unvaccinated individuals using a Cox regression-model, adjusted by prior SARS-CoV-2 infection status, vaccine-manufacturer/dosing-interval, demographics and workplace exposures.

**Results:** Of 35,768 participants, 27% (n=9,488) had a prior SARS-CoV-2 infection. Vaccine coverage was high: 97% had two-doses (79% BNT162b2 long-interval, 8% BNT162b2 short-interval, 8% ChAdOx1). There were 2,747 primary infections and 210 reinfections between 07/12/2020 and 21/09/2021. Adjusted VE (aVE) decreased from 81% (95% CI 68%-89%) 14-73 days after dose-2 to 46% (95% CI 22%-63%) >6-months; with no significant difference for short-interval BNT162b2 but significantly lower aVE (50% (95% CI 18%-70%) 14-73 days after dose-2 from ChAdOx1. Protection from infection-acquired immunity showed evidence of waning in unvaccinated follow-up but remained consistently over 90% in those who received two doses of vaccine, even in those infected over 15-months ago.

**Conclusion:** Two doses of BNT162b2 vaccination induce high short-term protection to SARS-CoV-2 infection, which wanes significantly after six months. Infection-acquired immunity boosted with vaccination remains high over a year after infection. Boosters will be essential to maintain protection in vaccinees who have not had primary infection to reduce infection and transmission in this population.

**Trial registration number:** ISRCTN11041050

## BACKGROUND

Understanding the durability of the immune response to SARS-CoV-2 infection and COVID-19 vaccination remains critical to the global COVID-19 response. Twenty months after emergence, SARS-CoV-2 has caused millions of deaths,^1^ and widespread disruption to global health and economies. The development and mass deployment of COVID-19 vaccines within a year was unprecedented and has facilitated relaxation of non-pharmaceutical interventions. COVID-19 vaccines have demonstrated short-term effectiveness in real-world studies, reducing both symptomatic and asymptomatic infection, severity and secondary transmission.^2-5^ The duration of this protection over longer periods remains uncertain and requires ongoing study.

Population uptake of COVID-19 vaccination in the UK (aged over 12 years) is 80.4% for two doses,^6^ and prioritised groups (health and social care workers and the clinically vulnerable), are now over six months after their second dose. Following concerns about potential vaccine waning at this point,^7-11^ and in the context of sustained high levels of community infections,^6^ the UK Government initiated a roll-out of booster vaccination to priority groups in September 2021.^12^ Improved understanding and characterisation of vaccine effectiveness at longer intervals and potential variation by demographic factors, vaccine schedules and history of SARS-CoV-2 infection is urgently required to support global vaccination schedules.

The SARS-CoV-2 Immunity and Reinfection Evaluation (SIREN) study, a large cohort of healthcare workers undergoing fortnightly asymptomatic PCR testing, had over 30% of participants testing seropositive at enrolment and is well suited to this task.^5,13,14^ In this analysis we estimate the effectiveness and durability of protection against future SARS-CoV-2 infection conferred by previous SARS-CoV-2 infection and COVID-19 vaccination in the SIREN cohort from March 2020 to September 2021.

## METHODS

### Study design and participants

The SIREN study is a multicentre prospective cohort of healthcare workers aged over 18 years across the UK.

### Data sources and measurement

Participants undergo fortnightly SARS-CoV-2 PCR testing (supplemented by widespread lateral flow testing), monthly antibody testing and complete regular questionnaires. This data collection is described elsewhere.^5^

Vaccination data (manufacturer, dates) were obtained via linkage on personal identifiers from national COVID-19 registries in each health administration and directly from participants in their questionnaires. Dosing interval was categorised as ‘short’ if dose-two was administered up to 6-weeks post dose-one and ‘long’ if ≥6-weeks.^15^

Serum samples from all participant baseline visits are collected centrally and tested at the United Kingdom Health Security Agency (UKHSA) central testing laboratory at Porton Down using the semi-quantitative Elecsys Anti-SARS-CoV-2 nucleocapsid (N) protein assay and fully quantitative Elecsys Anti-SARS-CoV-2 spike (S) protein assay (Roche Diagnostics).

### Outcomes

The primary outcome was a PCR-confirmed SARS-CoV-2 infection, irrespective of symptom status, that met the definition of a reinfection in the positive cohort of two PCR positives ≥90 days or one new PCR positive ≥28 days after an antibody positive result consistent with previous infection.

### Explanatory variables and exclusion criteria

Participants were assigned into one of two cohorts at the start of analysis time: participants in the naïve cohort had no history of SARS-CoV-2 positivity and the positive cohort being those who had ever received a PCR positive or antibody result consistent with prior SARS-CoV-2 infection.

Participants were excluded from this analysis if event or cohort assignment could not be accurately completed, i.e. no PCR tests during follow-up, or if they were in the positive cohort but were infected after vaccination or lacked an onset date for primary infection (PCR positive or COVID-symptom onset).

### Person time at risk

Follow-up began on 07 December 2020, the day before COVID-19 vaccination was introduced to the UK, and continued until 21 September 2021, covering 10 calendar months. All participants enrolled on or before 07 December 2020 contributed follow-up time from 07 December 2020 onwards. Participants enrolled after 07 December 2020 began contributing follow-up time from their enrolment date (delayed entry). Participants moved from the negative to positive cohort 90 days after a primary PCR positive date, if their primary infection was before vaccination, at which point they were considered at risk of reinfection (mirroring the SIREN reinfection definition: two PCR positives >90-days apart). End of follow-up time for individual participants was either date of primary infection (negative cohort), date of reinfection (positive cohort) or last PCR negative test.

### Statistical methods

We used a Cox proportional hazards model with delayed entry, the outcome being time-to-infection with a positive PCR test. The model accounted for the calendar time, varying infection rate via the baseline hazard, that could take any functional form. Analysis time started shortly before the second wave peaked, continuing through Spring 2021 and into the third wave (Supplementary Figure iii), thus, accounting for a varying hazard rate was crucial.

The main predictors – vaccine status and previous infection status - were categorical and time-varying. We grouped on the time to vaccination and divided follow-up time into unvaccinated and 24 post-vaccination time intervals, with post-vaccination intervals categorised by manufacturer, dose and dosing interval, the latter to explore differences in protection in those receiving dose two closer in time to their first dose. We also grouped the time since primary infection into four time-intervals: before primary infection (naïve), and then 3-9 months, 9-15 months and ≥15 months after primary infection. Vaccine effectiveness and protection from primary infection were calculated as 1-HR. We used robust variance estimates to guard against the potential for unmeasured confounders at trust level.

We initially fitted a main effects model, with no interactions between vaccine and primary infection status. This was our main model that highlighted vaccine effectiveness over time. We also fitted an interaction model, in which we did not consider time since vaccination, brand and manufacturer, to focus on protection from primary infection over time by vaccine status. We fitted both models with and without additional time invariant covariates: age, ethnicity, co-morbidities, region, frequency of COVID-19 patient contact, patient-facing role, and workplace setting. Independently, we also fitted an equivalent piecewise exponential proportional hazards model. This produced consistent VE results and provided estimates of the baseline hazard rates (supplementary material; Figure iii), analogous to the method we have previously described.^5^ We used STATA software (version 15.1; StataCorp LLC, College Station, TX, USA) for all analyses. Results were independently replicated in R (v. 4.1.1, survival package v.3.2-13). Our annotated code is available (https://github.com/SIREN-study/SARS-CoV-2-Immunity).

This study was registered, number ISRCTN11041050, and received approval from the Berkshire Research Ethics Committee on 22 May 2020. Reporting of the study follows the Strengthening the Reporting of Observational studies in Epidemiology guidelines.^16^

## RESULTS

### Study population

The SIREN study enrolled 44,546 participants between 18 June 2020 and 23 April 2021 from 135 sites across the UK; n=35,768 met the inclusion criteria for this analysis (Supplementary Figure i). Participants are described in Table 1, and were predominantly female (84%), with a median age of 46 years (IQR 36-54). We assigned 26,280 participants to the naïve cohort and 9,488 to the positive cohort at analysis start time. The positive cohort were more likely to be male, younger, from Black, Asian and ethnic minority backgrounds, work in clinical roles and report more frequent exposure to COVID-19 patients (Table 1). By the end of analysis time, 97% of the cohort had received two vaccine doses: 78.5% BNT162b2 long-interval, 8.6% BNT162b2 short-interval and 7.8% ChAdOX1 (Table 1, Supplementary Figure ii). We identified no major demographic differences between participants by vaccine schedule (Supplementary Table i). Follow-up time varied by participant, with a total of 7,482,388 participant person-days, of which there were 998,270 person-days unvaccinated, and 6,430,118 person-days vaccinated (from date of first dose). There were 60,301 PCR tests performed in the unvaccinated follow-up period and 443,979 PCR tests in the vaccinated follow-up period, with an average test interval of 16.6 days per test in the unvaccinated period and 14.5 days per test in the vaccinated period. There were 2,747 primary infections during follow-up and 210 reinfections, with cases peaking at the end of December 2020 and declining by March-April 2021, before increasing in May 2021, which mirrored national trends (Supplementary Figure iii).

**Table 1:**
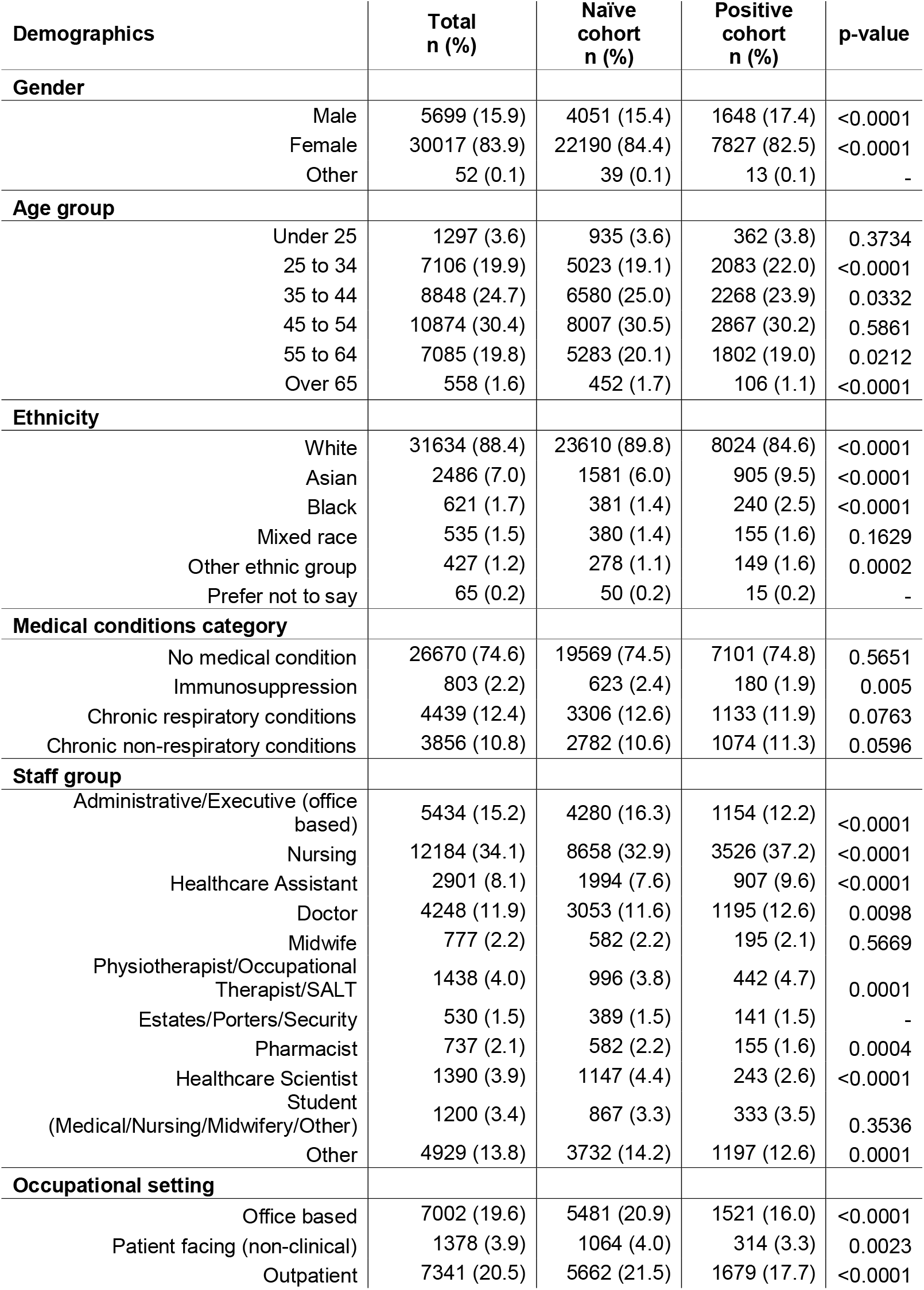
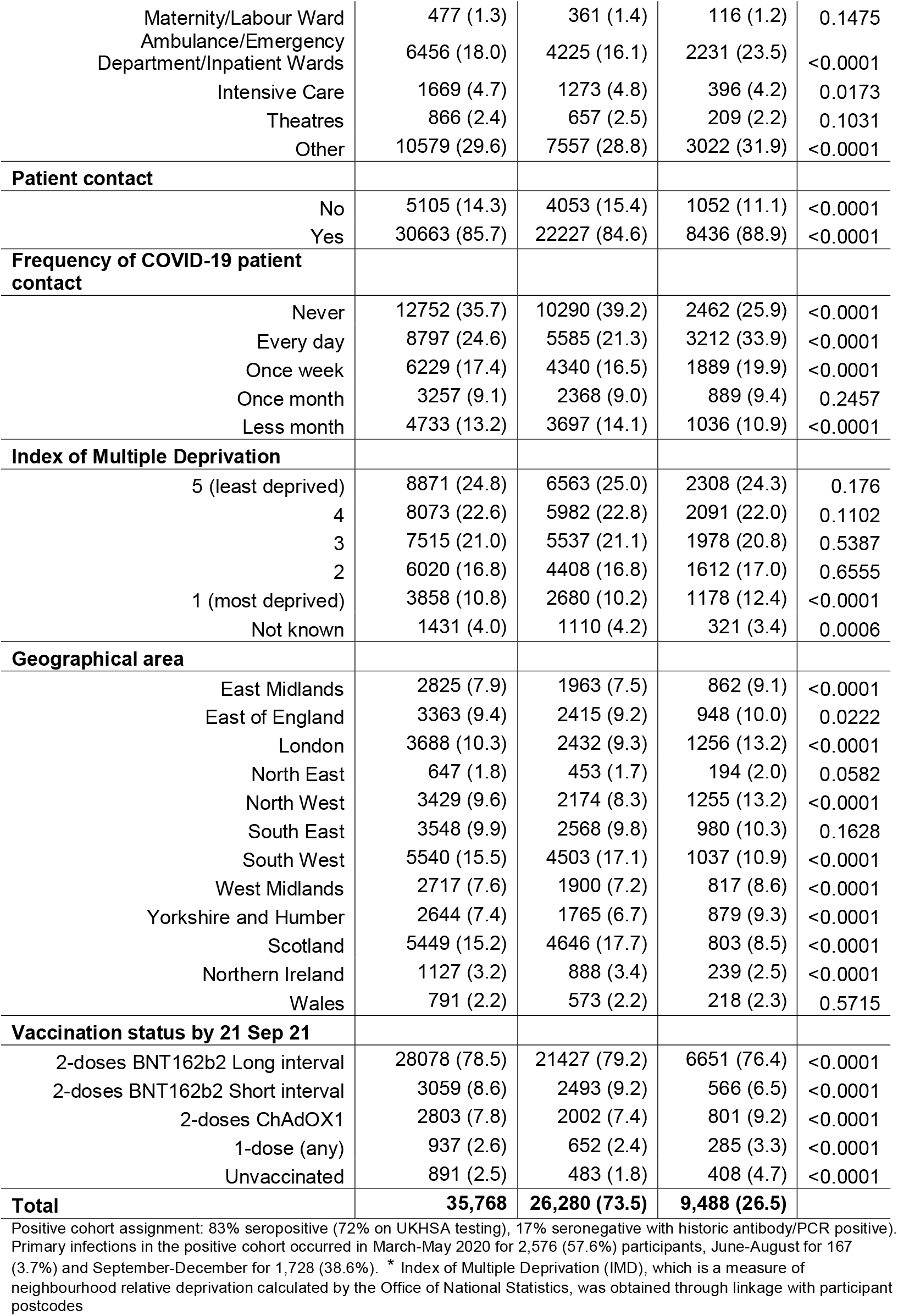
Description of participant demographics, by cohort assignment, June 2020 to September 2021.

### Vaccine effectiveness

The overall adjusted vaccine effectiveness (aVE) against infection following dose-two of BNT162b2 vaccine administered after the long-interval was 81% (95% CI 68%-89%) in the first two months after the development of the full immune response (14-73 days after second dose) (Table 2, Figure 1). aVE declined over time, although remained high at 70% (95% CI 62%-76%) 4-6 months after dose-two. After six months we saw evidence of waning, with aVE of 46% (95% CI 22%-63%).

**Table 2:** Incidence of SARS-CoV-2 infections and effectiveness of COVID-19 vaccines against infection by dose, manufacturer and dosing interval, SIREN participants 07 December 2020 to 21 September 2021.

| Vaccine status | Number of participants | Number of days of follow up | All infections (symptomatic & asymptomatic) |  |  |  |
| --- | --- | --- | --- | --- | --- | --- |
|  |  |  | Number of infections | Crude Incident rate (per 10,000) | VE (1-HR) 95% CI | aVE (1-HR) 95% CI |
| **Unvaccinated** | 24,787 | 995,122 | 1067 | 10.72 | Reference | Reference |
| **Vaccinated 1 dose** |  |  |  |  |  |  |
| Time since vaccine |  |  |  |  |  |  |
| *BNT162b2* |  |  |  |  |  |  |
| 21 – 27 days | 21,374 | 141,696 | 55 | 3.88 | 0.56 (0.40-0.68) | 0.57 (0.41-0.69) |
| 28 – 41 days | 21,137 | 279,672 | 62 | 2.22 | 0.62 (0.46-0.74) | 0.62 (0.46-0.74) |
| 42 – 55 days | 21,968 | 289,896 | 32 | 1.10 | 0.68 (0.52-0.78) | 0.67 (0.51-0.78) |
| > 55 days | 23,049 | 477,746 | 60 | 1.26 | 0.64 (0.51-0.73) | 0.58 (0.42-0.70) |
| *ChAdOX1* |  |  |  |  |  |  |
| 21 – 27 days | 2,299 | 15,848 | 3 | 1.89 | 0.46 (-0.82-0.84) | 0.42 (-0.92-0.83) |
| 28 – 41 days | 2,423 | 32,556 | 1 | 0.31 | 0.88 (0.17-0.98) | 0.87 (0.10-0.98) |
| 42 – 55 days | 2,488 | 33,514 | 3 | 0.90 | 0.46 (-0.56-0.81) | 0.40 (-0.75-0.80) |
| > 55 days | 2,503 | 64,708 | 10 | 1.55 | 0.36 (-0.30-0.68) | 0.29 (-0.43-0.65) |
| **Vaccinated 2 doses** |  |  |  |  |  |  |
| Time since vaccine |  |  |  |  |  |  |
| *BNT162b2 long-interval* |  |  |  |  |  |  |
| 14 – 73 days | 25,571 | 1,466,353 | 26 | 0.18 | 0.83 (0.70-0.90) | 0.81 (0.68-0.89) |
| 74 – 133 days | 23,776 | 1,297,486 | 285 | 2.20 | 0.70 (0.61-0.77) | 0.65 (0.56-0.73) |
| 134 – 193 days | 18,255 | 688,494 | 505 | 7.33 | 0.70 (0.61-0.77) | 0.67 (0.58-0.75) |
| >193 days | 2,704 | 24,575 | 83 | 33.77 | 0.43 (0.16-0.61) | 0.43 (0.17-0.61) |
| *BNT162b2 short-interval* |  |  |  |  |  |  |
| 14 – 73 days | 2,861 | 151,318 | 10 | 0.66 | 0.85 (0.70-0.92) | 0.85 (0.71-0.92) |
| 74 – 133 days | 2,822 | 164,199 | 6 | 0.37 | 0.72 (0.40-0.87) | 0.72 (0.42-0.86) |
| 134 – 193 days | 2,659 | 147,301 | 50 | 3.39 | 0.56 (0.38-0.69) | 0.55 (0.37-0.67) |
| >193 days | 2,105 | 84,705 | 90 | 10.63 | 0.59 (0.41-0.71) | 0.58 (0.40-0.71) |
| *ChAdOX1* |  |  |  |  |  |  |
| 14 – 73 days | 2,394 | 133,865 | 19 | 1.42 | 0.52 (0.20-0.71) | 0.49 (0.16-0.69) |
| 74 – 133 days | 2,003 | 92,621 | 55 | 5.94 | 0.53 (0.34-0.67) | 0.47 (0.26-0.63) |
| > 133 days | 995 | 23,226 | 32 | 13.78 | 0.59 (0.31-0.75) | 0.51 (0.18-0.71) |

Number of infections includes both primary infections and reinfections (all PCR confirmed)
Crude incident rate: number of infections/days of follow-up (\*10,000), does not adjust for variable baseline hazard.

**Figure 1:**
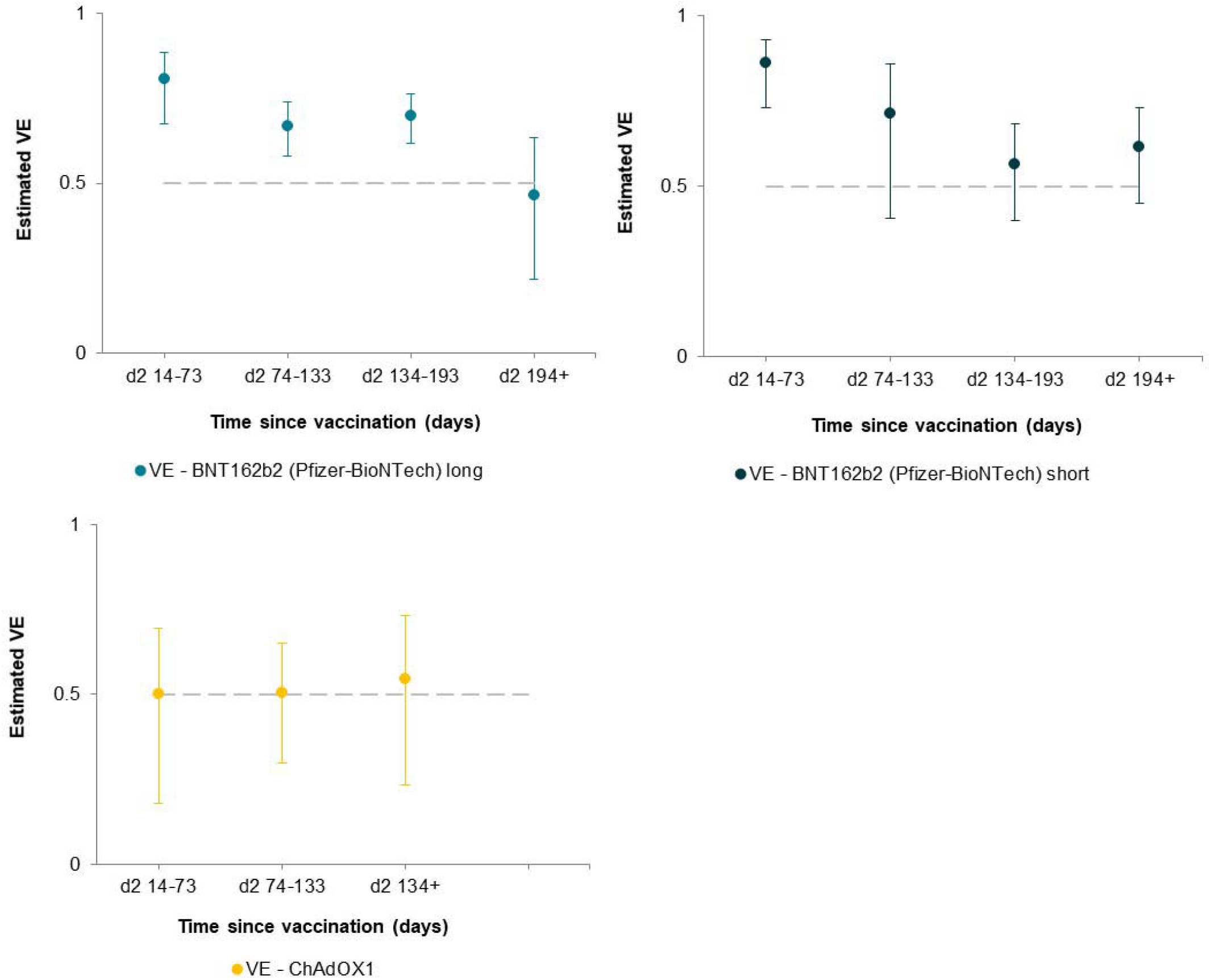
Adjusted Vaccine Effectiveness over time after two doses: BNT162b2 (Pfizer-BioNTech) short and long interval and ChAdOX1 (combined short and long interval) Number of participants: **BNT162b2 long-interval**: 14-73 days n=25571, 74-133 days n=23776, 134-193 days n=18255, over 193 days n=2704; **BNT162b2 short-interval**: 14-73 days n=2861, 74-133 days n=2822, 134-193 days n=2659, over 193 days n=2105; **ChAdOx1**: 14-73 days n=2394, 74-133 days n=2003, over 133 days n=995. aVE: adjusted Vaccine Effectiveness, model adjusted for time since vaccination and previous infection status (time since previous infection) and constant predictors: age, gender, ethnicity, comorbidities, workplace setting, frequency of contact with COVID-19 patients, geographical area (of workplace).

A similar trend was observed for BNT162b2 dose two short-interval, with higher protection at 14-73 days (aVE 86% (95% CI 73%-93%) decreasing to 61% (95% CI 45%-73%) after 6-months. We found no significant difference in protection after dose-two between BNT162b2 long and short inter-vaccination intervals, with HR for infection of 1.39 (95% CI 0.64-3.00, p=0.41) using short interval as the reference group.

For ChAdOX1, aVE from two doses was 50% (95% CI 16%-69%) 14-73 days after second dose. Effectiveness did not fall significantly after longer intervals after dose-two, with overlapping confidence intervals of VE reflecting the small number of participants contributing to this estimate (Table 2, Figure 1). Compared to ChAdOX1, we found that Pfizer short was 72% more effective (95% CI 38%-88%, *p*=0.002) and Pfizer long was 62% more effective (95% CI 26%-80%, *p*=0.004), in the interval 14-73 days.

### Durability of protection following primary infection

In contrast, looking at the impact of vaccination on the cohort with prior COVID-19 infection (positive cohort), using naïve unvaccinated as the reference group (Table 3, Figure 2), a beneficial boosting of infection-acquired immunity was apparent, with combined protection over 90% a year or more after primary infection and two doses of vaccination. There was no evidence of the protection afforded by primary infection waning in participants who had received two doses of vaccine up to 15 months after the primary infection. A similar trend was observed after a single dose and even without vaccination, however, most unvaccinated follow-up time occurred pre-Delta.

**Table 3:** Incidence of SARS-CoV-2 reinfections and durability of protection against SARS-CoV-2 reinfection, adjusted for vaccine status, in the SIREN cohort between 07 December 2020 and 21 September 2021.

| Vaccine and prior infection status | Number of participants | Number of days of follow up | Number of infections | Crude incident rate (per 10,000) | Adjusted Absolute protection against infection (1-HR) 95% CI |
| --- | --- | --- | --- | --- | --- |
| **Unvaccinated** |  |  |  |  |  |
| Naïve | 18,039 | 646,495 | 983 | 15.21 | Reference |
| Prior infection 3-9 months | 6,173 | 169,697 | 41 | 2.42 | 0.85 (0.80-0.89) |
| Prior infection 9-15 months | 4,145 | 163,437 | 32 | 1.96 | 0.85 (0.78-0.89) |
| Prior infection ≥15 months | 291 | 15,493 | 11 | 7.10 | 0.73 (0.43-0.87) |
| **Vaccinated dose 1** |  |  |  |  |  |
| Naïve | 21,283 | 1,273,056 | 607 | 4.77 | 0.35 (0.24-0.44) |
| Prior infection 3-9 months | 4,561 | 152,160 | 15 | 0.99 | 0.87 (0.79-0.92) |
| Prior infection 9-15 months | 5,978 | 358,618 | 24 | 0.67 | 0.90 (0.86-0.93) |
| Prior infection ≥15 months | 196 | 7,353 | 2 | 2.72 | 0.87 (0.50-0.97) |
| **Vaccinated dose 2** |  |  |  |  |  |
| Naïve | 22,586 | 3,414,257 | 1102 | 3.23 | 0.64 (0.56-0.70) |
| Prior infection 3-9 months | 2,928 | 320,252 | 24 | 0.75 | 0.91 (0.86-0.94) |
| Prior infection 9-15 months | 7,202 | 624,026 | 29 | 0.46 | 0.91 (0.86-0.94) |
| Prior infection ≥15 months | 4,980 | 280,388 | 30 | 1.07 | 0.95 (0.93-0.97) |

Number of infections includes both primary infections and reinfections (all PCR confirmed)
Crude incident rate: number of infections/days of follow-up (\*10,000), does not adjust for variable baseline hazard.

**Figure 2:**
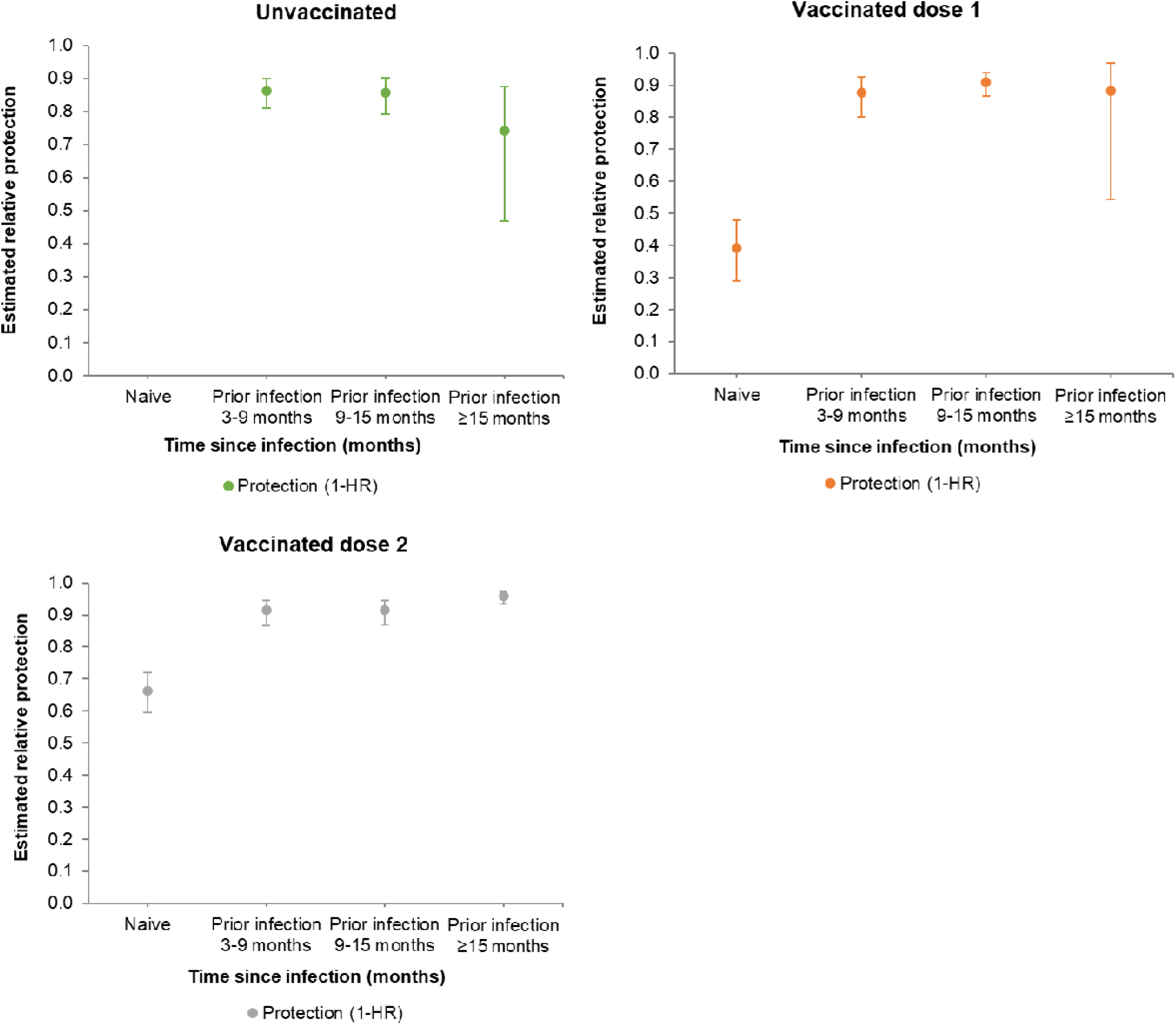
Protection following primary infection under different COVID-19 vaccination scenarios, up to 18 months following infection. Number of participants: **Unvaccinated:** prior infection 3-9 months n=6298, 9-15 months n=4147 and over 15 months n=291; **vaccinated dose 1**: naïve n=21283, prior infection 3-9 months n=4561, 9-15 months n=5978 and over 15 months n=196; **vaccinated dose 2**: naïve n=22586, prior infection 3-9 months n=2928, 9-15 months n=7202 and over 15 months n=4980. Adjusted absolute protection against infection: model adjusted for previous infection status (time since previous infection), vaccine status (unvaccinated, dose 1 (this includes follow-up between dose 1 and dose 2), dose 2 (includes follow-up after dose 2)) and constant predictors: age, gender, ethnicity, comorbidities, workplace setting, frequency of contact with COVID-19 patients, geographical area (of workplace). Reference group is unvaccinated infection naïve. The model does not adjust for time since vaccination (as in table 2), vaccine manufacturer or vaccine dosing interval.

## DISCUSSION

Eighteen months after the emergence of SARS-CoV-2 and ten months after the rapid deployment of COVID-19 vaccines, we have assessed the durability of protection from SARS-CoV-2 infection conferred by both infection-acquired and vaccine-acquired immunity. Our cohort of 35,768 healthcare workers, including over a quarter with prior infection, primarily received two doses of BNT162b2 administered at a long inter-vaccine interval, which induced high levels of protection over the first 6 months, peaking between 68% and 89% in the first two months; however, we found evidence of significant waning, with protection reducing to between 22% and 63% after six months. We found no difference in protection following two doses when comparing BNT162b2 short interval with BNT162b2 long interval, although we found significantly lower protection from two doses of ChAdOX1 compared to BNT12b2. Of note, the period of waning coincided with the Delta variant being the predominant circulating strain, which may account for the more pronounced waning of protection in our cohort, given the reduced vaccine effectiveness against Delta reported.^17^ Delivery of vaccination to individuals after prior infection effectively boosts and extends their immunity, with participants who received two doses of vaccination after infection emerging as the most highly protected group in our cohort for both symptomatic and asymptomatic infection, with similar protection to that provided by a three-course vaccination against symptomatic infection.^18^

Our finding of reduced protection from infection following two doses of vaccination after six months strengthens the accruing evidence base. Our design overcomes several biases of recent studies, including underestimation of the proportion with prior infection;^19^ previous studies have typically investigated symptomatic infection and utilised test-negative case-control or retrospective cohort designs using national testing surveillance data.^7,9,11^ We note that these real-world studies have found consistently lower protection and more pronounced waning than the recent BNT162b2 clinical trial, which reported vaccine efficacy against symptomatic infection of 83.7% (95% CI, 74.7 to 89.9) 4-6 months after dose-2,^20^ likely related to the reduced vaccine effectiveness reported against the Delta variant.^17^ The significantly lower protection observed in this study after ChAdOX1 compared to BNT162b2 has also been found in other recent studies.^7,20^ Several studies have observed lower antibody titres following ChAdOx1 vaccination than BNT162b2,^21,22^ and a shorter interval to fall below a protective antibody threshold from this lower baseline has been proposed as a causal mechanism for the lower vaccine effectiveness.^20^ We found no evidence of a difference in protection against infection after two doses of BNT162b2 between short and long-interval. This is despite evidence of significantly higher antibody, B cell and T cell responses in recipients of long-interval compared to short-interval vaccination regimens,^15,23,24^ and higher VE against symptomatic infection from one observational study.^15^ Plausibly the threshold to prevent all infections may be lower than that for symptomatic infection.

Studies to date have shown more durable protection against severe outcomes of hospitalisation and death following vaccination.^7,25^ Whilst we have estimated VE against all infections, including asymptomatic infections that have limited clinical impact, a reduction in VE against infection will increase transmission and risk of infection to high-risk individuals, some of whom will progress to severe disease.

To our knowledge, this study reports the longest real-world follow-up time from primary infection to date. It remains unclear how long immune protection will last after previous infection due to the limited length of follow-up period, however modelling has suggested that protection could last for up to 61 months.^21,26^ In our cohort, we have demonstrated that protection from primary infection can last up to 15 months in some individuals, while other studies have reported protection ranging from 5-12 months.^27-29^ Our ability to study infection-acquired immunity in unvaccinated individuals at longer intervals is now limited given the very small number of our cohort remaining unvaccinated. It is important to highlight that most follow-up without vaccination in the 9-15-month category occurred in the pre-Delta wave. It is possible that the sustained infection-acquired protection in our cohort is affected by repeated low dose occupational exposure to COVID-19,^30^ and therefore less generalisable to populations at lower exposure. It is also possible that this results from a broader diversity of T-cell immunity against different SARS-CoV-2 spike protein epitopes emerging following infection, enhancing protection against variants and inducing long-lasting memory T-cell populations.^27,31,32^ Although our finding of greater protection following infection-acquired immunity has been demonstrated by other authors,^33,34^ others have reported vaccine-acquired immunity to be equivalent,^35,36^ or superior.^37^ Despite the high protection provided by infection-acquired immunity, we have demonstrated additional benefit from vaccination in previously infected participants, in line with previous studies.^34,38,39^ Until thresholds for protective antibody titres against SARS-CoV-2 infection are established, it is challenging to accurately estimate how much vaccine-induced immunity is required to prevent reinfection at an individual level.

Key strengths of our study are the size of our cohort, asymptomatic testing and testing frequency, with an average PCR test interval of 16.6 days in unvaccinated time and 14.5 days per test in vaccinated follow-up time, supplemented by the widespread use of lateral flow testing, which means we can be confident that most infections were detected. As a well-defined cohort, we can simultaneously investigate vaccination and prior infection status and adjust for important confounders, including workplace exposures. The most important limitation of our study is the relatively small number of participants continuing to contribute follow-up time to key vaccination exposures: unvaccinated, ChAdOx1 and BNT162b2 short interval. This particularly affects the precision of estimates and our ability to assess potential waning following two-doses of ChAdOx1, and >15 months after primary infection in unvaccinated participants. We consider that the strengths of our study design and speed of vaccine deployment significantly limit the impact of depletion-of-susceptible bias (which particularly affects studies on vaccine-waning),^19^ however we recognise some residual confounding may remain.

## Conclusion

Two doses of BNT162b2 vaccination, given with a short or long-interval, induce high protection to SARS-CoV-2 infection (asymptomatic and symptomatic) in the short-term, but this protection wanes after six months, during a period where Delta predominates.

Protection provided from two doses of ChAdOX1 is considerably lower overall. The highest and most durable protection is observed in those with hybrid immunity, who received one or two doses of vaccine after a primary infection; this will be important for the deployment of vaccines in highly exposed and immune populations. Strategic use of booster vaccine doses to avert waning of protection (particularly in double vaccinated naïve individuals) is essential to provide reduced infection, and therefore transmission in the ongoing global response to COVID-19.

## Supporting information

Supplementary Figure iii

## Data Availability

Annotated code for this analysis is available at: (https://github.com/SIREN-study/SARS-CoV-2-Immunity). The metadata for this analysis will be available to researchers through the Health Data Research UK CO-CONNECT platform and available for secondary analysis.

## Acknowledgements

We would like to thank all participants for their ongoing contribution and commitment to this study, and to all the research teams for their hard-work and support delivering the study at all 135 sites. Thank you all for making SIREN possible.

We are grateful to colleagues at the UKHSA Seroepidemiology Unit for their support biobanking and processing the high-volumes of serum samples and to colleagues at UKHSA Porton Down who have organised and performed all the centralised serology testing, including testing 35,000 baseline samples, in particular Caoimhe Kelly, Anaya Ellis, Gabrielle Harker, Olivia Carr, Aaron Lloyd, Hannah Selman, Matthew Royds and Georgia Hemingway.

---

^□^**The SIREN study group**

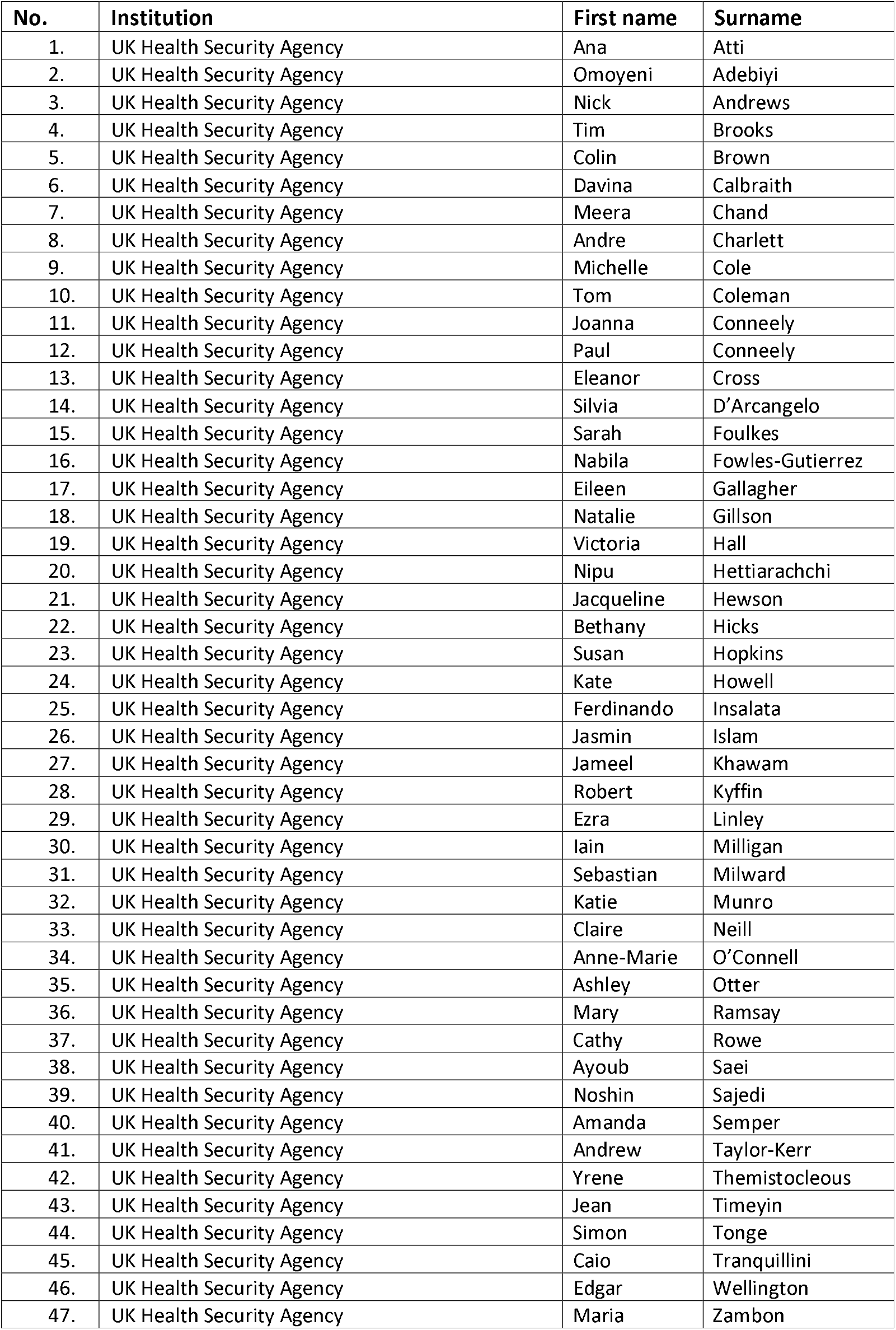
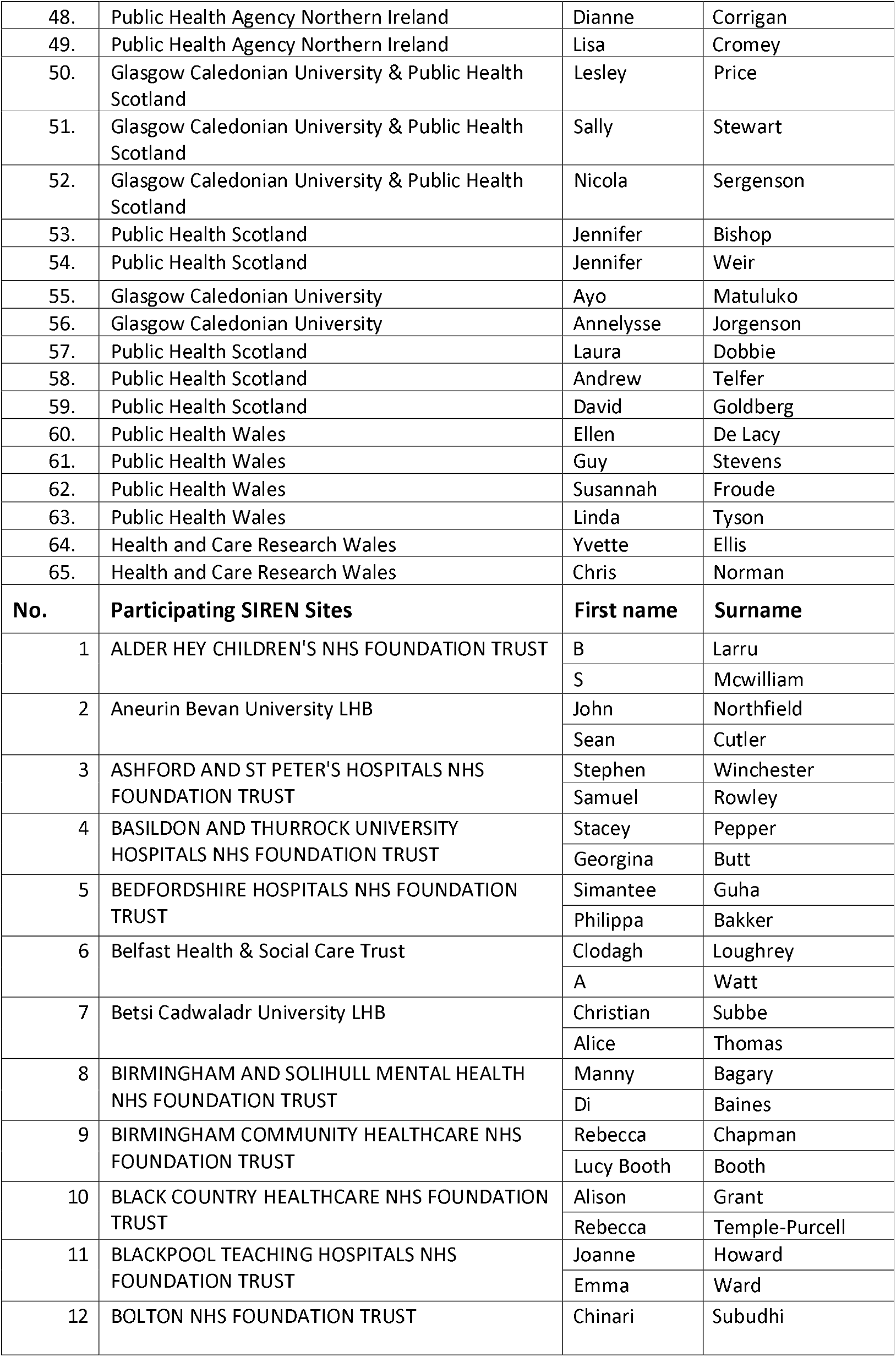
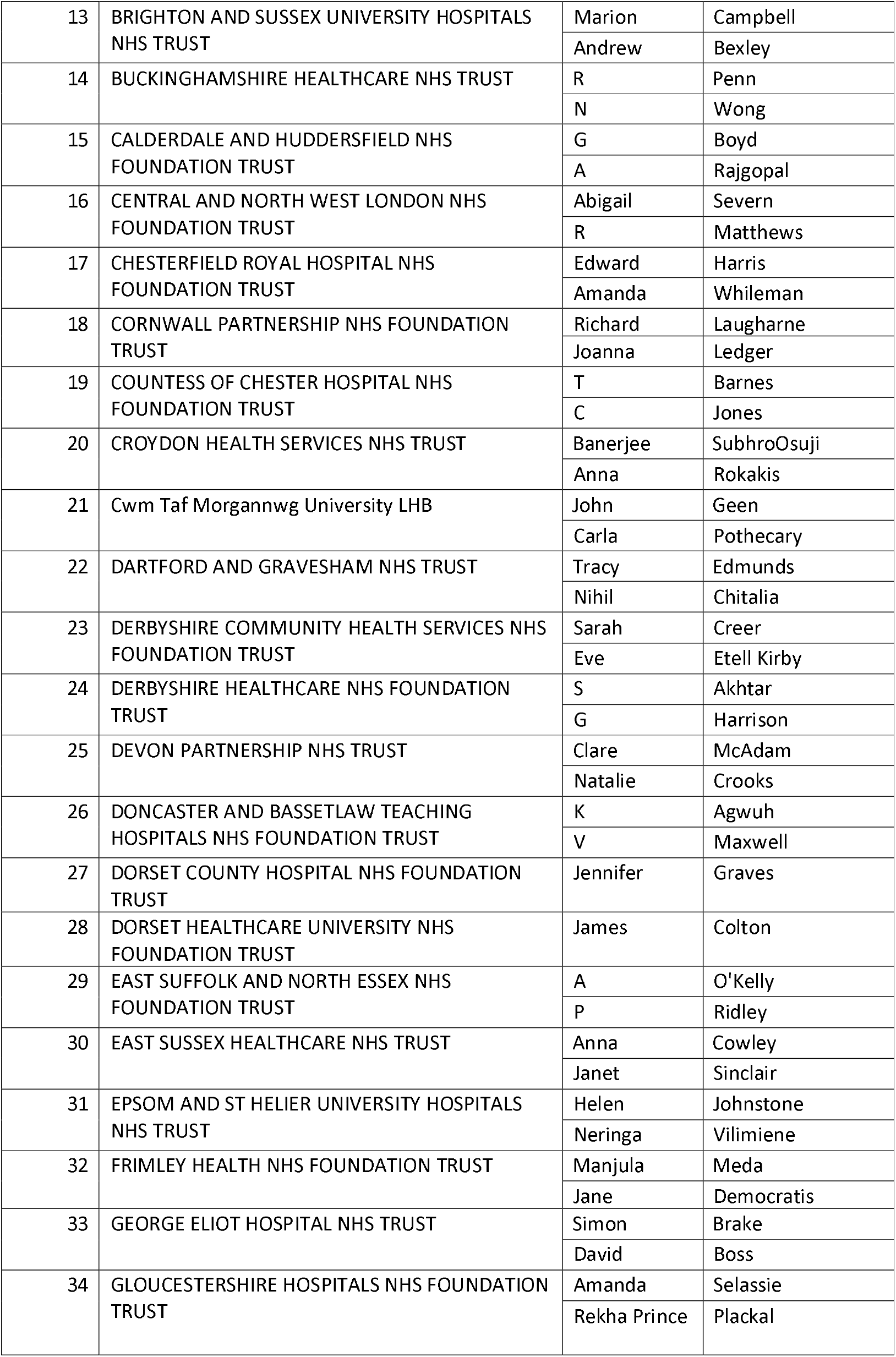
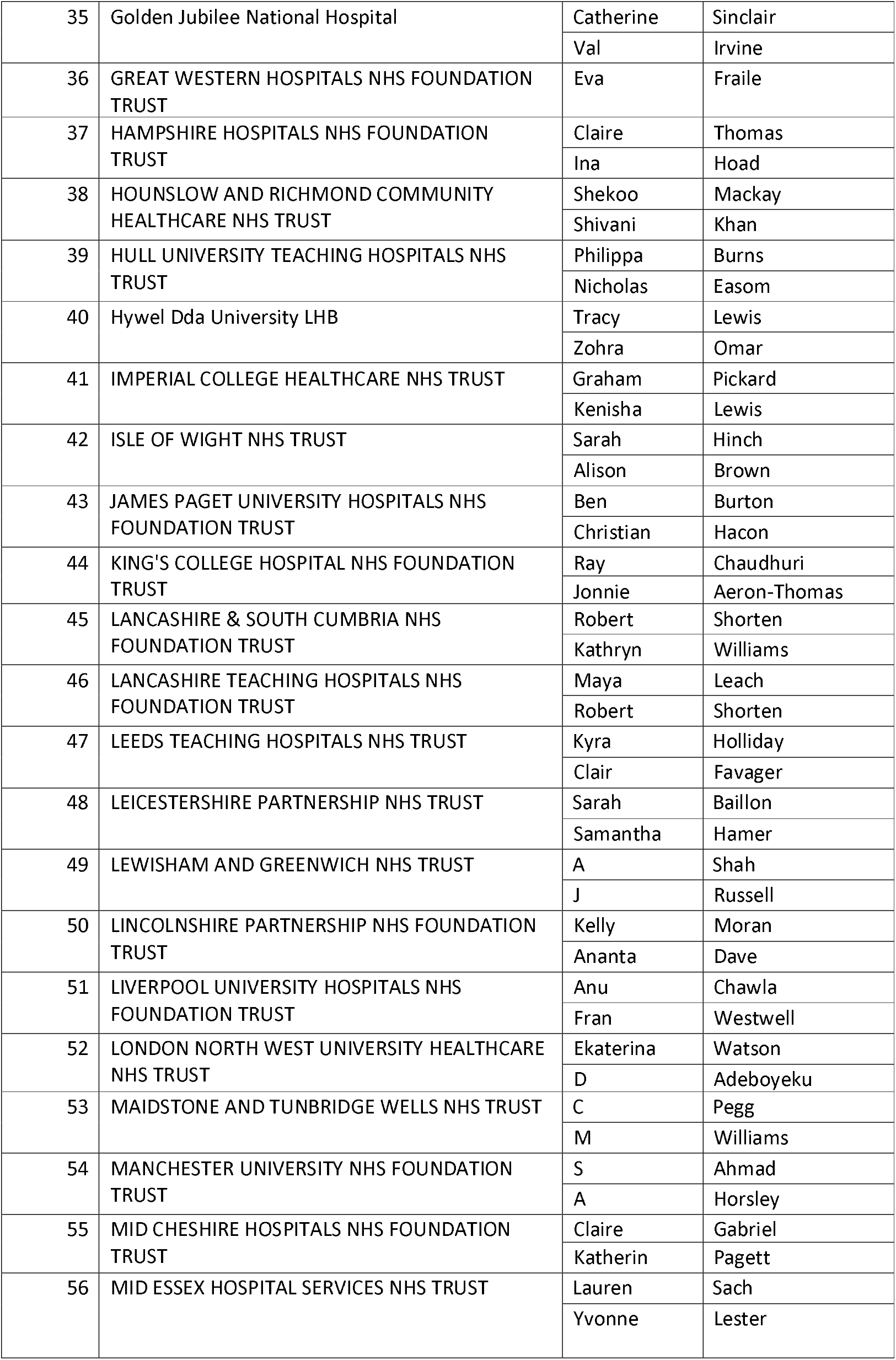
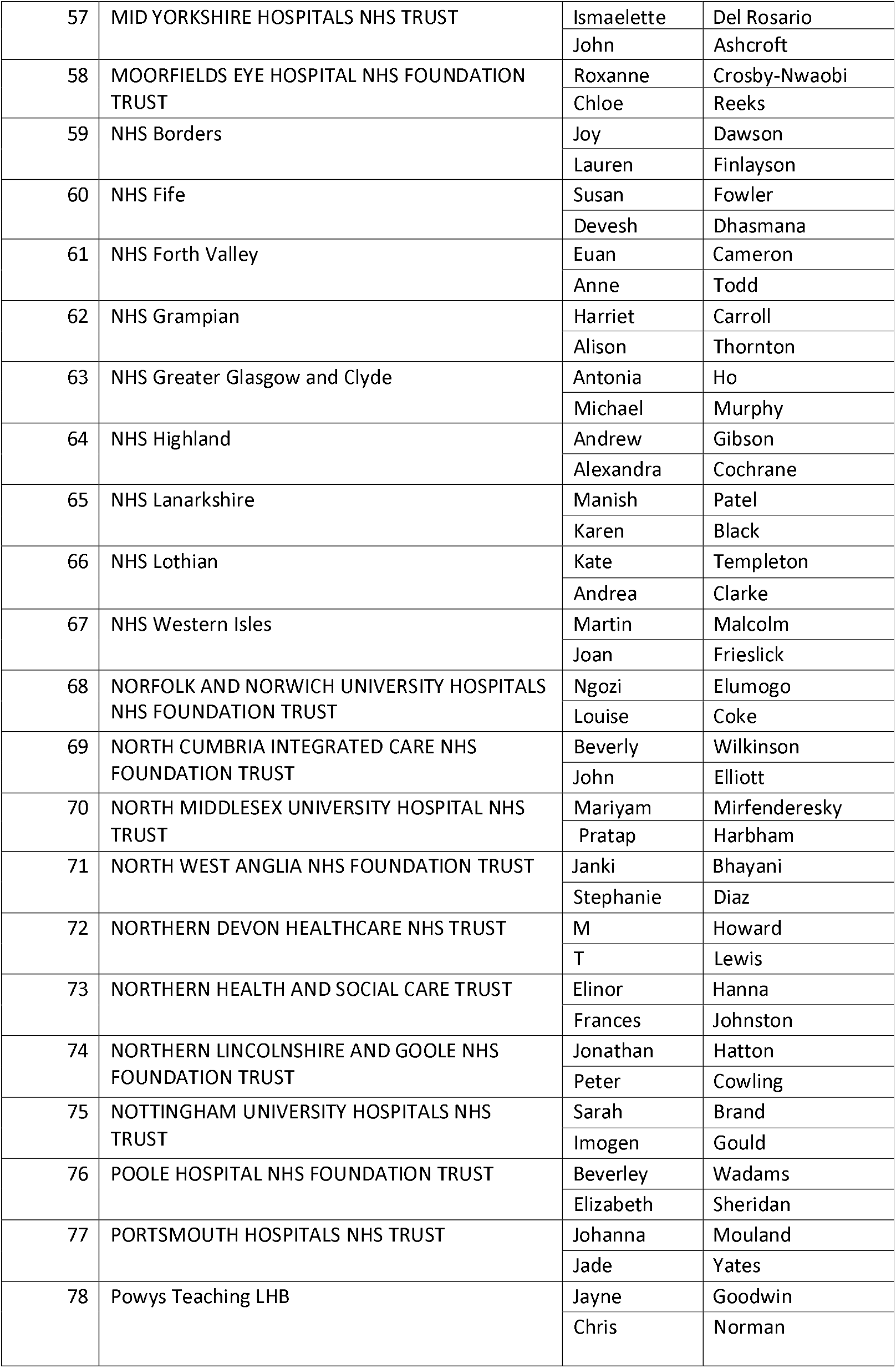
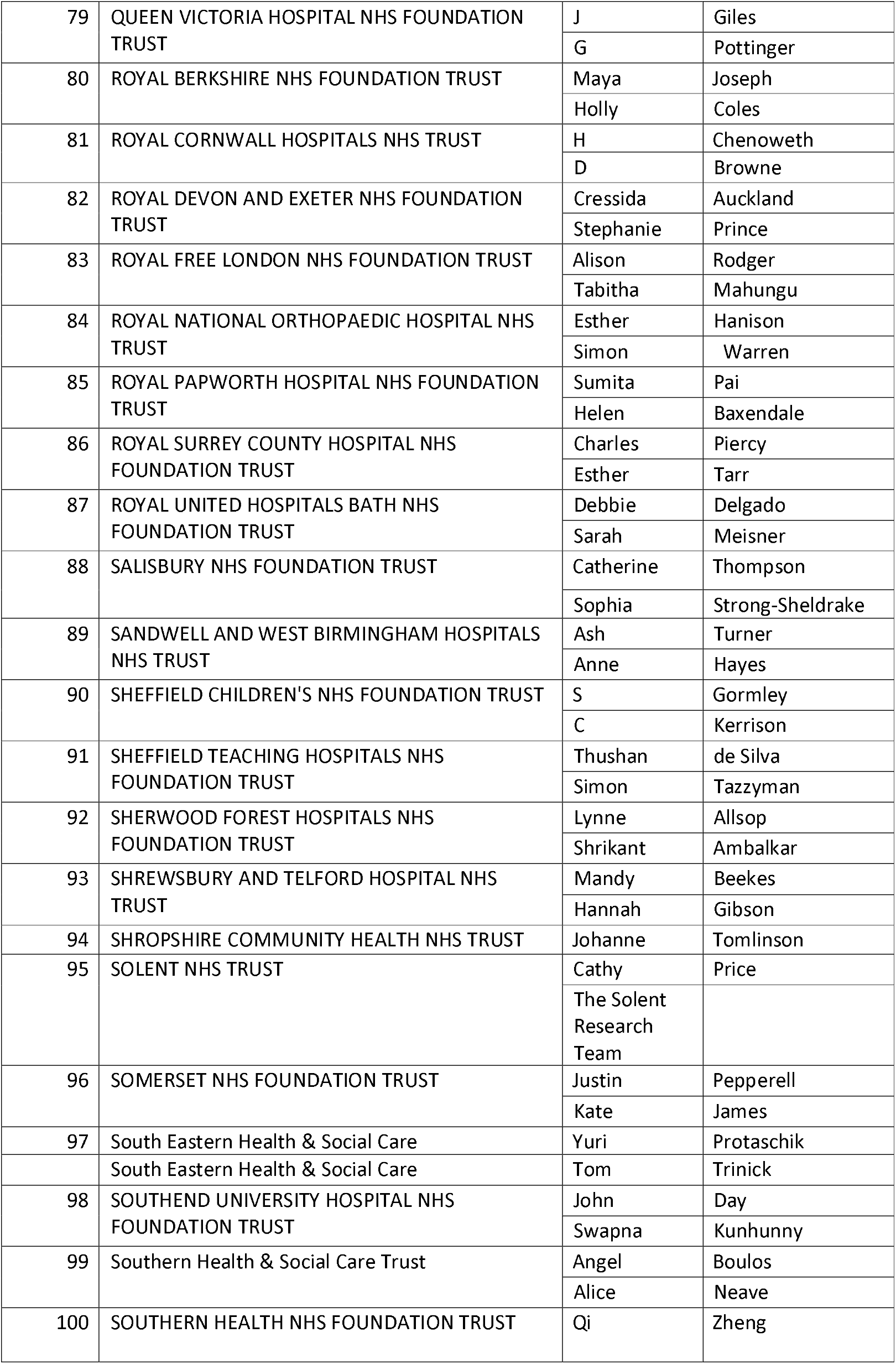
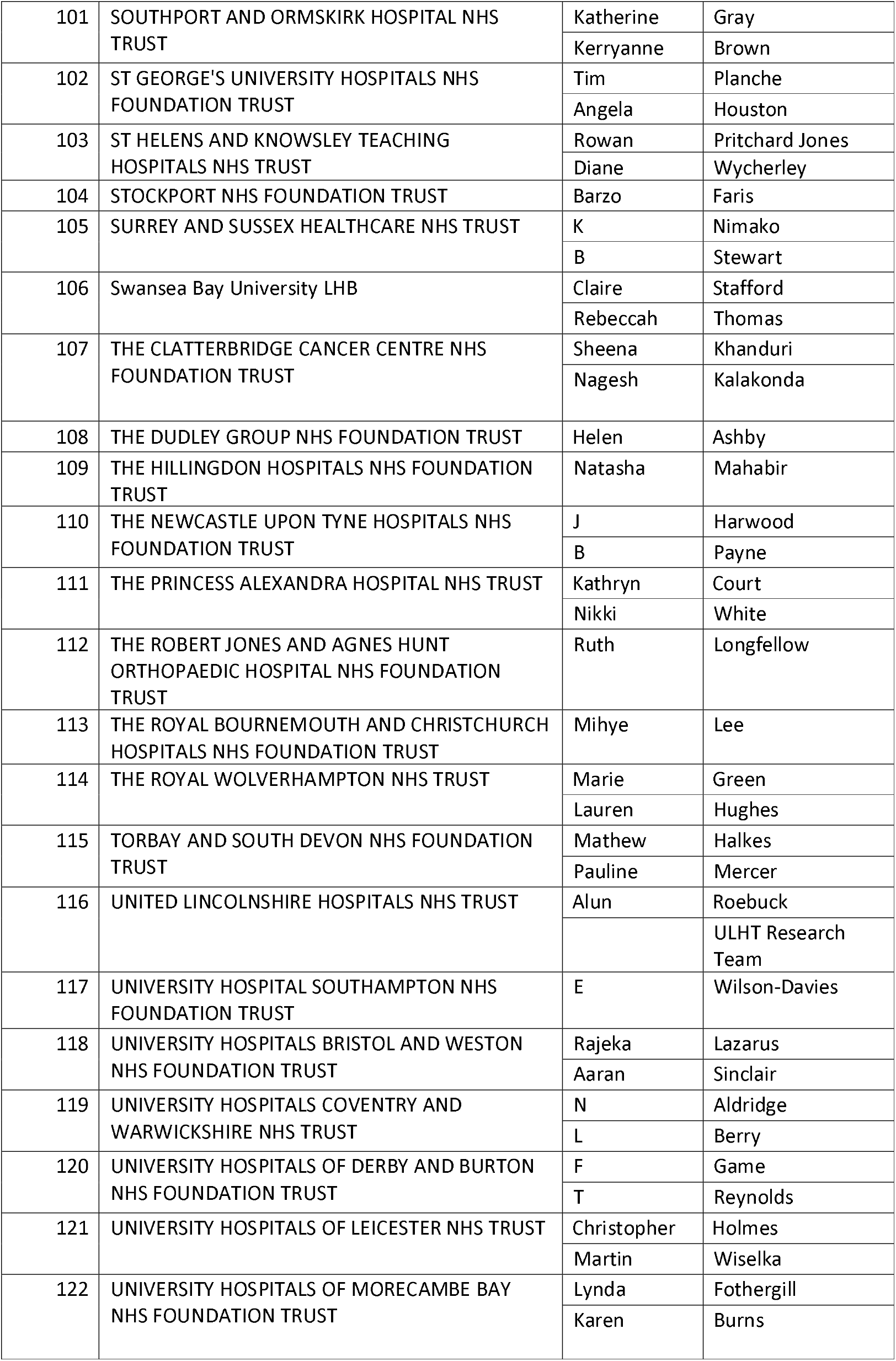
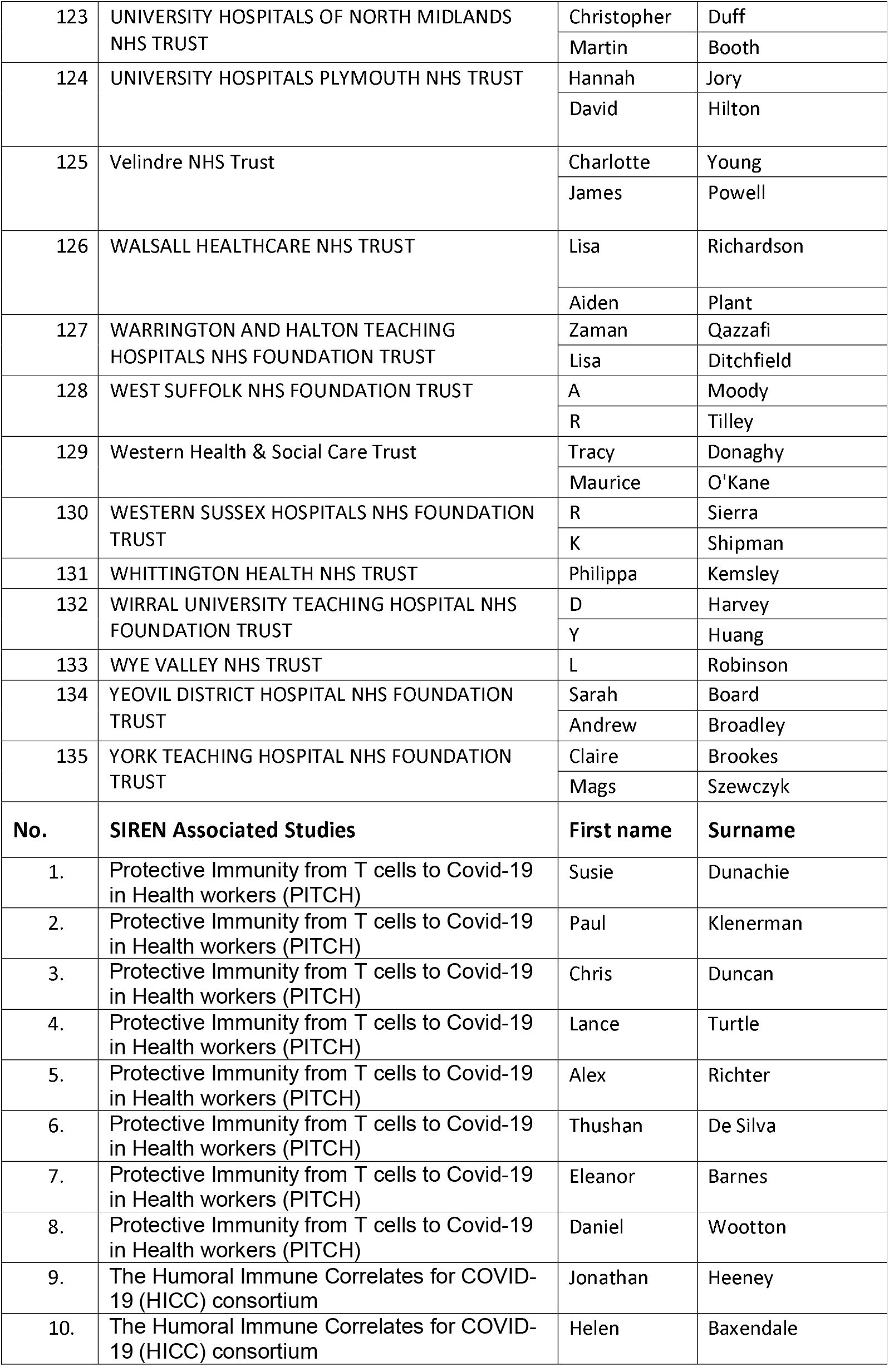
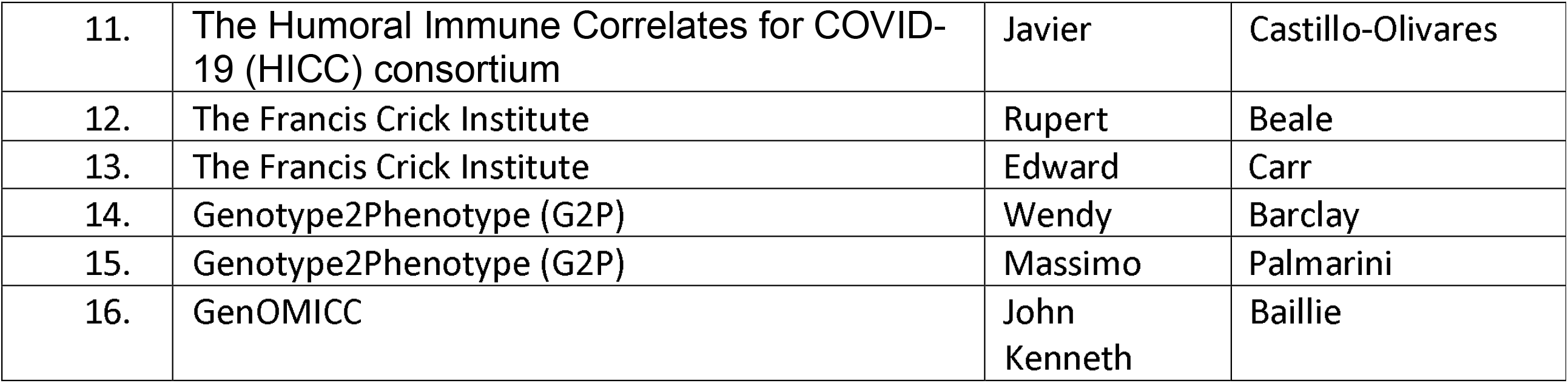

## Notes

### Competing Interest Statement

The authors have declared no competing interest.

### Clinical Protocols

https://www.medrxiv.org/content/10.1101/2020.12.15.20247981v1

### Funding Statement

The study is funded by the Department for Health and Social Care and UKHSA (formerly PHE) with contributions from the governments of Northern Ireland, Scotland and Wales. Funding was also provided by The National Institute for Health Research (NIHR) as an Urgent Public Health Priority Study and through the Health Protection Research Units. The study was also awarded grant funding from the Medical Research Council (Grant title: Investigation of proven vaccine breakthrough by SARS-CoV-2 variants in
established UK healthcare worker cohorts: SIREN consortium & PITCH Plus Pathway).

### Author Declarations

This study was registered, number ISRCTN11041050, and received approval from the Berkshire Research Ethics Committee on 22 May 2020.

