## Supplementary Figure i for "Effectiveness and durability of protection against future SARS-CoV-2 infection conferred by COVID-19 vaccination and previous infection; findings from the UK SIREN prospective cohort study of healthcare workers March 2020 to September 2021"

Supplementary Figure i: SIREN participant flow diagram

**296** withdrawals who requested their data to be removed

**265** excluded potential events of interest*

**18** unreliable vaccine data

**411** no PCR or antibody data

**3,494** cohort cannot be assigned

**100** unreliable antibody data

**4,194** did not meet inclusion criteria for the model

**44,546** participants enrolled into SIREN

**35,768** included in analysis

Events of interest excluded after initial screening include misreported results, false positive results, seroconversion between two RT-PCR positive results less than 90 days apart and seroconversion after vaccination confirmed by anti-N SARS-CoV-2 negativity.
