## Supplementary Table i for "Effectiveness and durability of protection against future SARS-CoV-2 infection conferred by COVID-19 vaccination and previous infection; findings from the UK SIREN prospective cohort study of healthcare workers March 2020 to September 2021"

**Supplementary Table i: description of participant demographics, by cohort assignment and vaccine status at the end of the analysis period, June 2020 to September 2021**

| **Demographics** | **Total** | **Naïve cohort** | | | | | | | **Positive cohort** | | | | | | |
| --- | --- | --- | --- | --- | --- | --- | --- | --- | --- | --- | --- | --- | --- | --- | --- |
|  |  | **Dose 2**  **n (%)** | | | | | **Unvaccinated**  **n (%)** | **P-value Pf long vs unvaccinated** | **Dose 2**  **n (%)** | | | | | **Unvaccinated**  **n (%)** | **P-value Pf long vs unvaccinated** |
|  |  | **Pfizer short** | **Pfizer long** | **P-value Pf long vs Pf short** | **ChAdOX1** | **P-value Pf long vs Ch** |  |  | **Pfizer short** | **Pfizer long** | **P-value Pf long vs Pf short** | **ChAdOX1** | **P-value Pf long vs Ch** |  |  |
| **Gender** |  |  |  |  |  |  |  |  |  |  |  |  |  |  |  |
| Male | 5699 (15.9) | 495 (20.2) | 3175 (15.2) | <0.0001 | 228 (12) | 0.0002 | 50 (10.8) | 0.0090 | 156 (25.6) | 1282 (17.7) | <0.0001 | 112 (12.3) | <0.0001 | 52 (12.1) | 0.0030 |
| Female | 30017 (83.9) | 1952 (79.7) | 17637 (84.6) | <0.0001 | 1666 (87.9) | 0.0001 | 410 (88.7) | 0.0155 | 453 (74.4) | 5942 (82.1) | <0.0001 | 796 (87.7) | <0.0001 | 375 (87.4) | 0.0051 |
| Other | 52 (0.1) | 3 (0.1) | 31 (0.1) | - | 1 (0.1) | - | 2 (0.4) | 0.0506 | - | 11 (0.2) | - | - | - | 2 (0.5) | 0.1943 |
| **Age group** |  |  |  |  |  |  |  |  |  |  |  |  |  |  |  |
| Under 25 | 1297 (3.6) | 58 (2.4) | 748 (3.6) | 0.0022 | 69 (3.6) | 1.0000 | 31 (6.7) | 0.0005 | 21 (3.4) | 285 (3.9) | 0.5385 | 35 (3.9) | 1.0000 | 12 (2.8) | 0.2493 |
| 25 to 34 | 7106 (19.9) | 379 (15.5) | 3920 (18.8) | <0.0001 | 391 (20.6) | 0.0556 | 167 (36.1) | <0.0001 | 126 (20.7) | 1531 (21.2) | 0.7717 | 184 (20.3) | 0.5310 | 144 (33.6) | <0.0001 |
| 35 to 44 | 8848 (24.7) | 639 (26.1) | 5158 (24.7) | 0.1293 | 485 (25.6) | 0.3849 | 119 (25.8) | 0.5878 | 153 (25.1) | 1710 (23.6) | 0.4033 | 219 (24.1) | 0.7383 | 105 (24.5) | 0.6700 |
| 45 to 54 | 10874 (30.4) | 781 (31.9) | 6410 (30.8) | 0.2651 | 564 (29.8) | 0.3663 | 90 (19.5) | <0.0001 | 194 (31.9) | 2213 (30.6) | 0.5042 | 284 (31.3) | 0.6664 | 95 (22.1) | 0.0002 |
| 55 to 64 | 7085 (19.8) | 533 (21.8) | 4255 (20.4) | 0.1047 | 357 (18.8) | 0.0972 | 50 (10.8) | <0.0001 | 106 (17.4) | 1419 (19.6) | 0.1876 | 175 (19.3) | 0.8299 | 67 (15.6) | 0.0417 |
| Over 65 | 558 (1.6) | 60 (2.4) | 352 (1.7) | 0.0130 | 29 (1.5) | 0.5170 | 5 (1.1) | 0.3219 | 9 (1.5) | 77 (1.1) | 0.3700 | 11 (1.2) | 0.7864 | 6 (1.4) | 0.5656 |
| **Ethnicity** |  |  |  |  |  |  |  |  |  |  |  |  |  |  |  |
| White | 31634 (88.4) | 2104 (85.9) | 18855 (90.5) | <0.0001 | 1697 (89.6) | 0.2024 | 386 (83.5) | <0.0001 | 492 (80.8) | 6174 (85.3) | 0.0029 | 772 (85) | 0.8100 | 345 (80.4) | 0.0057 |
| Asian | 2486 (7) | 228 (9.3) | 1204 (5.8) | <0.0001 | 96 (5.1) | 0.2098 | 28 (6.1) | 0.7851 | 81 (13.3) | 675 (9.3) | 0.0013 | 76 (8.4) | 0.3765 | 28 (6.5) | 0.0506 |
| Black | 621 (1.7) | 31 (1.3) | 262 (1.3) | - | 50 (2.6) | <0.0001 | 27 (5.8) | <0.0001 | 11 (1.8) | 152 (2.1) | 0.6181 | 34 (3.7) | 0.0023 | 34 (7.9) | <0.0001 |
| Mixed race | 535 (1.5) | 43 (1.8) | 284 (1.4) | 0.1162 | 30 (1.6) | 0.4806 | 11 (2.4) | 0.0725 | 5 (0.8) | 115 (1.6) | 0.1234 | 14 (1.5) | 0.8203 | 17 (4) | 0.0002 |
| Other ethnic group | 427 (1.2) | 40 (1.6) | 202 (1) | 0.0062 | 20 (1.1) | 0.6766 | 6 (1.3) | 0.5229 | 18 (3) | 110 (1.5) | 0.0048 | 10 (1.1) | 0.3428 | 4 (0.9) | 0.3152 |
| Prefer not to say | 65 (0.2) | 4 (0.2) | 36 (0.2) | 1.0000 | 2 (0.1) | 0.3406 | 4 (0.9) | 0.0013 | 2 (0.3) | 9 (0.1) | 0.1629 | 2 (0.2) | 0.3940 | 1 (0.2) | 0.5355 |
| **Medical conditions category** |  |  |  |  |  |  |  |  |  |  |  |  |  |  |  |
| No medical condition | 26670 (74.6) | 1747 (71.3) | 15616 (74.9) | 0.0001 | 1373 (72.5) | 0.0214 | 360 (77.9) | 0.1410 | 443 (72.7) | 5409 (74.8) | 0.2527 | 664 (73.1) | 0.2673 | 348 (81.1) | 0.0034 |
| Immunosuppression | 803 (2.2) | 88 (3.6) | 450 (2.2) | <0.0001 | 52 (2.7) | 0.1592 | 11 (2.4) | 0.7721 | 14 (2.3) | 135 (1.9) | 0.4909 | 16 (1.8) | 0.8347 | 12 (2.8) | 0.1903 |
| Chronic respiratory conditions | 4439 (12.4) | 337 (13.8) | 2577 (12.4) | 0.0478 | 262 (13.8) | - | 49 (10.6) | 0.2450 | 66 (10.8) | 871 (12) | 0.3799 | 116 (12.8) | 0.4858 | 36 (8.4) | 0.0247 |
| Chronic non-respiratory conditions | 3856 (10.8) | 278 (11.3) | 2200 (10.6) | 0.2885 | 208 (11) | 0.5886 | 42 (9.1) | 0.2996 | 86 (14.1) | 820 (11.3) | 0.0377 | 112 (12.3) | 0.3717 | 33 (7.7) | 0.0211 |
| **Staff group** |  |  |  |  |  |  |  |  |  |  |  |  |  |  |  |
| Administrative/Executive (office based) | 5434 (15.2) | 255 (10.4) | 3427 (16.4) | <0.0001 | 403 (21.3) | <0.0001 | 76 (16.5) | 0.9542 | 48 (7.9) | 902 (12.5) | 0.0008 | 131 (14.4) | 0.1053 | 48 (11.2) | 0.4278 |
| Nursing | 12184 (34.1) | 886 (36.2) | 6826 (32.7) | 0.0005 | 594 (31.3) | 0.2132 | 163 (35.3) | 0.2389 | 230 (37.8) | 2653 (36.7) | 0.5888 | 346 (38.1) | 0.4098 | 175 (40.8) | 0.0874 |
| Healthcare Assistant | 2901 (8.1) | 135 (5.5) | 1567 (7.5) | 0.0003 | 186 (9.8) | 0.0003 | 58 (12.6) | <0.0001 | 57 (9.4) | 671 (9.3) | 0.9350 | 89 (9.8) | 0.6258 | 56 (13.1) | 0.0092 |
| Doctor | 4248 (11.9) | 589 (24) | 2287 (11) | <0.0001 | 86 (4.5) | <0.0001 | 25 (5.4) | 0.0001 | 143 (23.5) | 910 (12.6) | <0.0001 | 72 (7.9) | <0.0001 | 35 (8.2) | 0.0071 |
| Midwife | 777 (2.2) | 46 (1.9) | 457 (2.2) | 0.3349 | 52 (2.7) | 0.1592 | 16 (3.5) | 0.0612 | 9 (1.5) | 152 (2.1) | 0.3160 | 20 (2.2) | 0.8434 | 10 (2.3) | 0.7795 |
| Physiotherapist/Occupational Therapist/SALT | 1438 (4) | 71 (2.9) | 816 (3.9) | 0.0143 | 66 (3.5) | 0.3872 | 13 (2.8) | 0.2257 | 19 (3.1) | 348 (4.8) | 0.0562 | 45 (5) | 0.7909 | 15 (3.5) | 0.2177 |
| Estates/Porters/Security | 530 (1.5) | 27 (1.1) | 304 (1.5) | 0.1182 | 30 (1.6) | 0.7324 | 14 (3) | 0.0094 | 7 (1.1) | 100 (1.4) | 0.5418 | 20 (2.2) | 0.0607 | 7 (1.6) | 0.7329 |
| Pharmacist | 737 (2.1) | 70 (2.9) | 463 (2.2) | 0.0279 | 29 (1.5) | 0.0439 | 4 (0.9) | 0.0580 | 14 (2.3) | 111 (1.5) | 0.1263 | 15 (1.7) | 0.6427 | 9 (2.1) | 0.3258 |
| Healthcare Scientist | 1390 (3.9) | 42 (1.7) | 978 (4.7) | <0.0001 | 74 (3.9) | 0.1127 | 20 (4.3) | 0.6876 | 6 (1) | 191 (2.6) | 0.0147 | 24 (2.6) | 1.0000 | 13 (3) | 0.6145 |
| Student (Medical/Nursing/Midwifery/Other) | 1200 (3.4) | 41 (1.7) | 709 (3.4) | <0.0001 | 81 (4.3) | 0.0406 | 17 (3.7) | 0.7251 | 16 (2.6) | 259 (3.6) | 0.1986 | 37 (4.1) | 0.4492 | 11 (2.6) | 0.2764 |
| Other | 4929 (13.8) | 288 (11.8) | 3009 (14.4) | 0.0005 | 294 (15.5) | 0.1928 | 56 (12.1) | 0.1631 | 60 (9.9) | 938 (13) | 0.0277 | 109 (12) | 0.3966 | 50 (11.7) | 0.4355 |
| **Occupational setting** |  |  |  |  |  |  |  |  |  |  |  |  |  |  |  |
| Office based | 7002 (19.6) | 363 (14.8) | 4402 (21.1) | <0.0001 | 501 (26.4) | <0.0001 | 77 (16.7) | 0.0217 | 80 (13.1) | 1208 (16.7) | 0.0213 | 149 (16.4) | 0.8192 | 47 (11) | 0.0019 |
| Patient facing (non-clinical) | 1378 (3.9) | 102 (4.2) | 829 (4) | 0.6336 | 81 (4.3) | 0.5247 | 19 (4.1) | 0.9136 | 19 (3.1) | 240 (3.3) | 0.7903 | 26 (2.9) | 0.5221 | 17 (4) | 0.4330 |
| Outpatient | 7341 (20.5) | 527 (21.5) | 4524 (21.7) | 0.8202 | 365 (19.3) | 0.0149 | 111 (24) | 0.2359 | 96 (15.8) | 1266 (17.5) | 0.2876 | 165 (18.2) | 0.6014 | 88 (20.5) | 0.1135 |
| Maternity/Labour Ward | 477 (1.3) | 37 (1.5) | 282 (1.4) | 0.6913 | 24 (1.3) | 0.7220 | 8 (1.7) | 0.5881 | 13 (2.1) | 80 (1.1) | 0.0281 | 11 (1.2) | 0.7864 | 7 (1.6) | 0.3407 |
| Ambulance/Emergency Department/Inpatient Wards | 6456 (18) | 495 (20.2) | 3309 (15.9) | <0.0001 | 225 (11.9) | <0.0001 | 85 (18.4) | 0.1467 | 154 (25.3) | 1685 (23.3) | 0.2633 | 191 (21) | 0.1209 | 115 (26.8) | 0.0967 |
| Intensive Care | 1669 (4.7) | 179 (7.3) | 962 (4.6) | <0.0001 | 73 (3.9) | 0.1612 | 28 (6.1) | 0.1292 | 30 (4.9) | 306 (4.2) | 0.4111 | 36 (4) | 0.7765 | 15 (3.5) | 0.4806 |
| Theatres | 866 (2.4) | 101 (4.1) | 506 (2.4) | <0.0001 | 29 (1.5) | 0.0128 | 8 (1.7) | 0.3294 | 26 (4.3) | 150 (2.1) | 0.0005 | 19 (2.1) | 1.0000 | 5 (1.2) | 0.2012 |
| Other | 10579 (29.6) | 646 (26.4) | 6029 (28.9) | 0.0096 | 597 (31.5) | 0.0171 | 126 (27.3) | 0.4529 | 191 (31.4) | 2300 (31.8) | 0.8387 | 311 (34.3) | 0.1282 | 135 (31.5) | 0.8968 |
| **Patient contact** |  |  |  |  |  |  |  |  |  |  |  |  |  |  |  |
| No | 5105 (14.3) | 209 (8.5) | 3311 (15.9) | <0.0001 | 350 (18.5) | 0.0032 | 59 (12.8) | 0.0710 | 54 (8.9) | 807 (11.2) | 0.0818 | 122 (13.4) | 0.0497 | 40 (9.3) | 0.2235 |
| Yes | 30663 (85.7) | 2241 (91.5) | 17532 (84.1) | <0.0001 | 1545 (81.5) | 0.0032 | 403 (87.2) | 0.0710 | 555 (91.1) | 6428 (88.8) | 0.0818 | 786 (86.6) | 0.0497 | 389 (90.7) | 0.2235 |
| **Frequency of COVID-19 patient contact** |  |  |  |  |  |  |  |  |  |  |  |  |  |  |  |
| Never | 12752 (35.7) | 674 (27.5) | 8363 (40.1) | <0.0001 | 852 (45) | <0.0001 | 145 (31.4) | 0.0002 | 118 (19.4) | 1896 (26.2) | 0.0002 | 273 (30.1) | 0.0122 | 104 (24.2) | 0.3594 |
| Every day | 8797 (24.6) | 706 (28.8) | 4218 (20.2) | <0.0001 | 394 (20.8) | 0.5338 | 135 (29.2) | <0.0001 | 227 (37.3) | 2435 (33.7) | 0.0717 | 282 (31.1) | 0.1175 | 158 (36.8) | 0.1875 |
| Once week | 6229 (17.4) | 511 (20.9) | 3396 (16.3) | <0.0001 | 252 (13.3) | 0.0007 | 79 (17.1) | 0.6453 | 150 (24.6) | 1430 (19.8) | 0.0046 | 161 (17.7) | 0.1327 | 83 (19.3) | 0.8006 |
| Once month | 3257 (9.1) | 234 (9.6) | 1903 (9.1) | 0.4169 | 143 (7.5) | 0.0196 | 42 (9.1) | 1.0000 | 59 (9.7) | 671 (9.3) | 0.7445 | 82 (9) | 0.7689 | 41 (9.6) | 0.8355 |
| Less month | 4733 (13.2) | 325 (13.3) | 2963 (14.2) | 0.2260 | 254 (13.4) | 0.3385 | 61 (13.2) | 0.5422 | 55 (9) | 803 (11.1) | 0.1108 | 110 (12.1) | 0.3680 | 43 (10) | 0.4800 |
| **Index of Multiple Deprivation** |  |  |  |  |  |  |  |  |  |  |  |  |  |  |  |
| 5 (least deprived) | 8871 (24.8) | 672 (27.4) | 5350 (25.7) | 0.0692 | 383 (20.2) | <0.0001 | 72 (15.6) | <0.0001 | 162 (26.6) | 1823 (25.2) | 0.4454 | 194 (21.4) | 0.0124 | 82 (19.1) | 0.0045 |
| 4 | 8073 (22.6) | 569 (23.2) | 4771 (22.9) | 0.7383 | 473 (25) | 0.0378 | 82 (17.7) | 0.0084 | 147 (24.1) | 1633 (22.6) | 0.3962 | 179 (19.7) | 0.0478 | 86 (20) | 0.2099 |
| 3 | 7515 (21) | 505 (20.6) | 4482 (21.5) | 0.3042 | 403 (21.3) | 0.8392 | 78 (16.9) | 0.0171 | 123 (20.2) | 1501 (20.7) | 0.7698 | 229 (25.2) | 0.0018 | 74 (17.2) | 0.0811 |
| 2 | 6020 (16.8) | 363 (14.8) | 3532 (16.9) | 0.0084 | 362 (19.1) | 0.0148 | 84 (18.2) | 0.4611 | 95 (15.6) | 1225 (16.9) | 0.4099 | 165 (18.2) | 0.3262 | 74 (17.2) | 0.8721 |
| 1 (most deprived) | 3858 (10.8) | 238 (9.7) | 2037 (9.8) | 0.8748 | 245 (12.9) | <0.0001 | 97 (21) | <0.0001 | 63 (10.3) | 875 (12.1) | 0.1887 | 130 (14.3) | 0.0575 | 77 (17.9) | 0.0004 |
| Not known | 1431 (4) | 103 (4.2) | 671 (3.2) | 0.0088 | 29 (1.5) | <0.0001 | 49 (10.6) | <0.0001 | 19 (3.1) | 178 (2.5) | 0.3667 | 11 (1.2) | 0.0149 | 36 (8.4) | <0.0001 |
| **Geographical area** |  |  |  |  |  |  |  |  |  |  |  |  |  |  |  |
| East Midlands | 2825 (7.9) | 263 (10.7) | 1308 (6.3) | <0.0001 | 335 (17.7) | <0.0001 | 28 (6.1) | 0.8610 | 61 (10) | 606 (8.4) | 0.1745 | 138 (15.2) | <0.0001 | 34 (7.9) | 0.7164 |
| East of England | 3363 (9.4) | 127 (5.2) | 2056 (9.9) | <0.0001 | 168 (8.9) | 0.1613 | 23 (5) | 0.0005 | 33 (5.4) | 793 (11) | <0.0001 | 62 (6.8) | 0.0001 | 40 (9.3) | 0.2724 |
| London | 3688 (10.3) | 283 (11.6) | 1913 (9.2) | 0.0001 | 127 (6.7) | 0.0003 | 62 (13.4) | 0.0021 | 115 (18.9) | 944 (13) | <0.0001 | 98 (10.8) | 0.0611 | 65 (15.2) | 0.1898 |
| North East | 647 (1.8) | 84 (3.4) | 333 (1.6) | <0.0001 | 26 (1.4) | 0.5043 | 3 (0.6) | 0.0881 | 20 (3.3) | 146 (2) | 0.0317 | 14 (1.5) | 0.3037 | 9 (2.1) | 0.8859 |
| North West | 3429 (9.6) | 281 (11.5) | 1432 (6.9) | <0.0001 | 371 (19.6) | <0.0001 | 52 (11.3) | 0.0002 | 85 (14) | 882 (12.2) | 0.1946 | 193 (21.3) | <0.0001 | 58 (13.5) | 0.4253 |
| South East | 3548 (9.9) | 260 (10.6) | 2139 (10.3) | 0.6444 | 112 (5.9) | <0.0001 | 23 (5) | 0.0002 | 83 (13.6) | 782 (10.8) | 0.0341 | 63 (6.9) | 0.0003 | 28 (6.5) | 0.0049 |
| South West | 5540 (15.5) | 285 (11.6) | 3843 (18.4) | <0.0001 | 246 (13) | <0.0001 | 58 (12.6) | 0.0014 | 58 (9.5) | 820 (11.3) | 0.1755 | 87 (9.6) | 0.1245 | 37 (8.6) | 0.0843 |
| West Midlands | 2717 (7.6) | 111 (4.5) | 1563 (7.5) | <0.0001 | 158 (8.3) | 0.2074 | 41 (8.9) | 0.2594 | 24 (3.9) | 630 (8.7) | <0.0001 | 102 (11.2) | 0.0130 | 39 (9.1) | 0.7754 |
| Yorkshire and Humber | 2644 (7.4) | 149 (6.1) | 1420 (6.8) | 0.1907 | 144 (7.6) | 0.1874 | 31 (6.7) | 0.9327 | 42 (6.9) | 703 (9.7) | 0.0235 | 80 (8.8) | 0.3856 | 43 (10) | 0.8385 |
| Scotland | 5449 (15.2) | 428 (17.5) | 3908 (18.7) | 0.1485 | 173 (9.1) | <0.0001 | 91 (19.7) | 0.5858 | 50 (8.2) | 645 (8.9) | 0.5591 | 56 (6.2) | 0.0062 | 37 (8.6) | 0.8320 |
| Northern Ireland | 1127 (3.2) | 78 (3.2) | 487 (2.3) | 0.0059 | 7 (0.4) | <0.0001 | 48 (10.4) | <0.0001 | 10 (1.6) | 121 (1.7) | 0.8542 | 4 (0.4) | 0.0029 | 29 (6.8) | <0.0001 |
| Wales | 791 (2.2) | 101 (4.1) | 441 (2.1) | <0.0001 | 28 (1.5) | 0.0776 | 2 (0.4) | 0.0110 | 28 (4.6) | 163 (2.3) | 0.0005 | 11 (1.2) | 0.0323 | 10 (2.3) | 1.0000 |
| **Total** | **35768** | **2450** | **20843** |  | **1895** |  | **462** |  | **609** | **7235** |  | **908** |  | **429** |  |

*Index of Multiple Deprivation (IMD), which is a measure of neighbourhood relative deprivation calculated by the Office of National Statistics, was obtained through linkage with participant postcodes; For ease of presentation, we have excluded participants who only received dose-1 (n=630 in the native cohort and n=307 in the positive cohort). Please note that this table presents and compares the demographics for the small number of participants who remained unvaccinated at the end of the analysis period, however, most participants contributed unvaccinated follow-up time to this analysis.
