## Supplementary Table ii for "Effectiveness and durability of protection against future SARS-CoV-2 infection conferred by COVID-19 vaccination and previous infection; findings from the UK SIREN prospective cohort study of healthcare workers March 2020 to September 2021"

Supplementary Table ii: Incidence of SARS-CoV-2 reinfections and durability of protection against SARS-CoV-2 reinfection, adjusted for vaccine status, in the SIREN cohort between 07 December 2020 and 21 September 2021 – full model

|  | **Hazard ratio** | **95% confidence interval** | |
| --- | --- | --- | --- |
|  |  | **Lower** | **High** |
| **Vaccine status** |  |  |  |
| Unvaccinated | - | - | - |
| d1 0-20 PF | 0.71 | 0.61 | 0.83 |
| d1 21-27 PF | 0.40 | 0.29 | 0.56 |
| d1 28-41 PF | 0.35 | 0.24 | 0.51 |
| d1 42-55 PF | 0.31 | 0.21 | 0.46 |
| d1 56+ PF | 0.40 | 0.29 | 0.55 |
| d1 0-20 AZ | 0.67 | 0.42 | 1.05 |
| d1 21-27 AZ | 0.54 | 0.16 | 1.81 |
| d1 28-41 AZ | 0.13 | 0.02 | 0.85 |
| d1 42-55 AZ | 0.57 | 0.20 | 1.68 |
| d1 56+ AZ | 0.69 | 0.34 | 1.39 |
| d2 0-13 PF long | 0.20 | 0.10 | 0.40 |
| d2 14-73 PF long | 0.19 | 0.11 | 0.32 |
| d2 74-133 PF long | 0.33 | 0.26 | 0.42 |
| d2 134-193 PF long | 0.30 | 0.24 | 0.38 |
| d2 194+ PF long | 0.54 | 0.37 | 0.78 |
| d2 0-13 PF short | 0.31 | 0.18 | 0.54 |
| d2 14-73 PF short | 0.14 | 0.07 | 0.27 |
| d2 74-133 PF short | 0.29 | 0.14 | 0.59 |
| d2 134-193 PF short | 0.44 | 0.32 | 0.60 |
| d2 194+ PF short | 0.39 | 0.27 | 0.55 |
| d2 14-73 AZ | 0.50 | 0.30 | 0.82 |
| d2 74-133 AZ | 0.50 | 0.35 | 0.70 |
| d2 134+ AZ | 0.45 | 0.27 | 0.77 |
| **Primary** **infection** |  |  |  |
| Naive | - | - | - |
| 3-9 months | 0.17 | 0.13 | 0.22 |
| 9-15 months | 0.17 | 0.14 | 0.21 |
| 15+ months | 0.15 | 0.10 | 0.21 |
| **Geography area** |  |  |  |
| East Midlands | - | - | - |
| East of England | 1.25 | 0.92 | 1.70 |
| London | 1.20 | 1.02 | 1.42 |
| North East | 1.06 | 0.74 | 1.51 |
| North West | 0.98 | 0.79 | 1.20 |
| South East | 0.87 | 0.70 | 1.07 |
| South West | 0.86 | 0.70 | 1.05 |
| West Midlands | 1.12 | 0.88 | 1.42 |
| Yorkshire and the Humber | 0.86 | 0.71 | 1.03 |
| Scotland | 0.88 | 0.68 | 1.15 |
| Northern Ireland | 0.77 | 0.49 | 1.21 |
| Wales | 1.52 | 0.99 | 2.33 |
| **Age group** |  |  |  |
| Under 25 | - | - | - |
| 25 to 34 | 0.72 | 0.60 | 0.86 |
| 35 to 44 | 0.61 | 0.51 | 0.72 |
| 45 to 54 | 0.57 | 0.48 | 0.68 |
| 55 to 64 | 0.49 | 0.42 | 0.57 |
| Over 65 | 0.33 | 0.22 | 0.48 |
| **Gender** |  |  |  |
| Male | - | - | - |
| Female | 0.95 | 0.84 | 1.07 |
| Non-binary | 0.46 | 0.06 | 3.32 |
| Prefer not to say | 0.45 | 0.06 | 3.40 |
| **Ethnicity groups** |  |  |  |
| White | - | - | - |
| Mixed Race | 0.98 | 0.77 | 1.24 |
| Asian | 1.20 | 1.03 | 1.40 |
| Black | 1.56 | 1.22 | 1.99 |
| Other ethnic group | 0.87 | 0.63 | 1.20 |
| Prefer not to say | 1.16 | 0.52 | 2.59 |
| **Work exposure frequency** |  |  |  |
| Never | - | - | - |
| Every day | 1.63 | 1.46 | 1.81 |
| Once week | 1.35 | 1.20 | 1.52 |
| Once month | 1.31 | 1.15 | 1.49 |
| Less month | 1.25 | 1.12 | 1.40 |
| **Occupational setting** |  |  |  |
| Office | - | - | - |
| Patient facing (non-clinical) | 0.99 | 0.80 | 1.24 |
| Outpatient | 1.08 | 0.95 | 1.23 |
| Maternity/Labour Ward | 0.93 | 0.64 | 1.37 |
| Ambulance/Emergency Department/Inpatient Wards | 1.38 | 1.19 | 1.61 |
| Intensive Care | 0.85 | 0.69 | 1.04 |
| Theatres | 0.91 | 0.71 | 1.17 |
| Other | 1.09 | 0.94 | 1.25 |
