## Supplementary Figure iv for "Effectiveness and durability of protection against future SARS-CoV-2 infection conferred by COVID-19 vaccination and previous infection; findings from the UK SIREN prospective cohort study of healthcare workers March 2020 to September 2021"

Supplementary Figure iv: Baseline hazard rates (for infection), estimated by a piecewise exponential model equivalent to the Cox model used to estimate protection from vaccine and primary infection.

| Date | Rate of infection |
| --- | --- |
| 2020-12-08 | 1 |
| 2020-12-09 | 0.583488 |
| 2020-12-10 | 1.16869 |
| 2020-12-11 | 0.418891 |
| 2020-12-12 | 0.42029 |
| 2020-12-13 | 0.758078 |
| 2020-12-14 | 1.9841 |
| 2020-12-15 | 1.73764 |
| 2020-12-16 | 1.48796 |
| 2020-12-17 | 1.23938 |
| 2020-12-18 | 1.50148 |
| 2020-12-19 | 0.862353 |
| 2020-12-20 | 0.605968 |
| 2020-12-21 | 1.69578 |
| 2020-12-22 | 1.39921 |
| 2020-12-23 | 1.58277 |
| 2020-12-24 | 1.1961 |
| 2020-12-25 | 0.2675 |
| 2020-12-26 | 1.20428 |
| 2020-12-27 | 1.65439 |
| 2020-12-28 | 1.43625 |
| 2020-12-29 | 3.07478 |
| 2020-12-30 | 3.0316 |
| 2020-12-31 | 1.73382 |
| 2021-01-01 | 0.618834 |
| 2021-01-02 | 1.15255 |
| 2021-01-03 | 1.30551 |
| 2021-01-04 | 2.77964 |
| 2021-01-05 | 1.8312 |
| 2021-01-06 | 1.41175 |
| 2021-01-07 | 1.64356 |
| 2021-01-08 | 1.36224 |
| 2021-01-09 | 0.960998 |
| 2021-01-10 | 1.40782 |
| 2021-01-11 | 2.47296 |
| 2021-01-12 | 2.24743 |
| 2021-01-13 | 1.8461 |
| 2021-01-14 | 2.06175 |
| 2021-01-15 | 1.49376 |
| 2021-01-16 | 1.27309 |
| 2021-01-17 | 0.982219 |
| 2021-01-18 | 1.55768 |
| 2021-01-19 | 1.59536 |
| 2021-01-20 | 1.44489 |
| 2021-01-21 | 1.81051 |
| 2021-01-22 | 1.35922 |
| 2021-01-23 | 1.10055 |
| 2021-01-24 | 1.0406 |
| 2021-01-25 | 1.97053 |
| 2021-01-26 | 1.36721 |
| 2021-01-27 | 1.54976 |
| 2021-01-28 | 1.13393 |
| 2021-01-29 | 1.31397 |
| 2021-01-30 | 0.313423 |
| 2021-02-01 | 0.292216 |
| 2021-02-02 | 1.33496 |
| 2021-02-03 | 0.512685 |
| 2021-02-04 | 0.785552 |
| 2021-02-05 | 0.711427 |
| 2021-02-06 | 0.271377 |
| 2021-02-07 | 0.183425 |
| 2021-02-08 | 0.937839 |
| 2021-02-09 | 0.85735 |
| 2021-02-10 | 0.386101 |
| 2021-02-11 | 0.684113 |
| 2021-02-12 | 0.494882 |
| 2021-02-13 | 0.698361 |
| 2021-02-14 | 0.200623 |
| 2021-02-15 | 0.405934 |
| 2021-02-16 | 0.713213 |
| 2021-02-17 | 0.409541 |
| 2021-02-18 | 0.102866 |
| 2021-02-19 | 0.518086 |
| 2021-02-20 | 0.104073 |
| 2021-02-21 | 0.208635 |
| 2021-02-23 | 0.437263 |
| 2021-02-24 | 0.520993 |
| 2021-02-25 | 0.728316 |
| 2021-02-26 | 0.103857 |
| 2021-02-27 | 0.103722 |
| 2021-02-28 | 0.104058 |
| 2021-03-01 | 0.310937 |
| 2021-03-02 | 0.30839 |
| 2021-03-04 | 0.0528012 |
| 2021-03-05 | 0.402914 |
| 2021-03-06 | 0.10049 |
| 2021-03-07 | 0.100537 |
| 2021-03-09 | 0.051515 |
| 2021-03-10 | 0.590697 |
| 2021-03-11 | 0.392577 |
| 2021-03-12 | 0.195794 |
| 2021-03-13 | 0.390692 |
| 2021-03-14 | 0.0975683 |
| 2021-03-15 | 0.0970695 |
| 2021-03-16 | 0.0968053 |
| 2021-03-17 | 0.389276 |
| 2021-03-18 | 0.391145 |
| 2021-03-20 | 0.100485 |
| 2021-03-21 | 0.0982346 |
| 2021-03-23 | 0.152747 |
| 2021-03-24 | 0.0998417 |
| 2021-03-25 | 0.101161 |
| 2021-03-27 | 0.10532 |
| 2021-03-29 | 0.160848 |
| 2021-03-30 | 0.3195 |
| 2021-03-31 | 0.107644 |
| 2021-04-01 | 0.432129 |
| 2021-04-04 | 0.0758231 |
| 2021-04-06 | 0.114433 |
| 2021-04-07 | 0.341882 |
| 2021-04-08 | 0.115441 |
| 2021-04-13 | 0.0506966 |
| 2021-04-14 | 0.119863 |
| 2021-04-15 | 0.241247 |
| 2021-04-16 | 0.121401 |
| 2021-04-19 | 0.0413315 |
| 2021-04-23 | 0.0627765 |
| 2021-04-28 | 0.0250046 |
| 2021-05-05 | 0.0178962 |
| 2021-05-06 | 0.119583 |
| 2021-05-07 | 0.118847 |
| 2021-05-13 | 0.0200743 |
| 2021-05-17 | 0.028994 |
| 2021-05-24 | 0.0164657 |
| 2021-05-29 | 0.0216776 |
| 2021-06-01 | 0.0342113 |
| 2021-06-02 | 0.200076 |
| 2021-06-04 | 0.0502188 |
| 2021-06-05 | 0.0979495 |
| 2021-06-06 | 0.194014 |
| 2021-06-07 | 0.192236 |
| 2021-06-08 | 0.477696 |
| 2021-06-09 | 0.0951838 |
| 2021-06-10 | 0.0950183 |
| 2021-06-12 | 0.0951258 |
| 2021-06-13 | 0.0933244 |
| 2021-06-14 | 0.371515 |
| 2021-06-16 | 0.139954 |
| 2021-06-17 | 0.185583 |
| 2021-06-18 | 0.0927912 |
| 2021-06-19 | 0.462806 |
| 2021-06-20 | 0.184566 |
| 2021-06-21 | 0.460094 |
| 2021-06-22 | 0.368622 |
| 2021-06-23 | 0.554633 |
| 2021-06-24 | 0.556656 |
| 2021-06-26 | 0.140223 |
| 2021-06-27 | 0.558141 |
| 2021-06-28 | 0.83758 |
| 2021-06-29 | 0.466828 |
| 2021-06-30 | 1.31305 |
| 2021-07-01 | 1.03688 |
| 2021-07-02 | 0.663206 |
| 2021-07-03 | 0.380832 |
| 2021-07-04 | 0.286174 |
| 2021-07-05 | 0.764189 |
| 2021-07-06 | 0.960161 |
| 2021-07-07 | 0.483378 |
| 2021-07-08 | 1.16961 |
| 2021-07-09 | 1.17908 |
| 2021-07-10 | 1.28445 |
| 2021-07-11 | 0.792579 |
| 2021-07-12 | 1.49058 |
| 2021-07-13 | 1.30233 |
| 2021-07-14 | 1.31351 |
| 2021-07-15 | 3.16265 |
| 2021-07-16 | 1.44335 |
| 2021-07-17 | 0.935623 |
| 2021-07-18 | 1.5671 |
| 2021-07-19 | 2.51679 |
| 2021-07-20 | 2.01171 |
| 2021-07-21 | 1.28562 |
| 2021-07-22 | 0.97324 |
| 2021-07-23 | 0.545943 |
| 2021-07-24 | 0.661199 |
| 2021-07-25 | 0.331675 |
| 2021-07-26 | 0.997096 |
| 2021-07-27 | 1.45613 |
| 2021-07-28 | 0.793739 |
| 2021-07-29 | 1.83461 |
| 2021-07-30 | 0.812083 |
| 2021-07-31 | 0.936473 |
| 2021-08-01 | 0.704446 |
| 2021-08-02 | 1.2958 |
| 2021-08-03 | 1.07272 |
| 2021-08-04 | 1.32685 |
| 2021-08-05 | 1.34281 |
| 2021-08-06 | 1.48192 |
| 2021-08-07 | 0.996316 |
| 2021-08-08 | 0.87478 |
| 2021-08-09 | 1.50506 |
| 2021-08-10 | 2.78859 |
| 2021-08-11 | 1.66707 |
| 2021-08-12 | 2.46883 |
| 2021-08-13 | 1.57747 |
| 2021-08-14 | 1.7218 |
| 2021-08-15 | 2.12699 |
| 2021-08-16 | 2.53705 |
| 2021-08-17 | 1.62312 |
| 2021-08-18 | 2.74413 |

The baseline hazard is relative to a female participant, white ethnicity, unvaccinated and with no previous infection, aged 45 to 54, working in an East Midlands site in an office setting with no exposure to patients. Estimates have been plotted until the 17 September 2021, as later estimates come with high uncertainty. Data at https://github.com/SIREN-study/SARS-CoV-2-Immunity.
