## Supplementary Figure iii for "Effectiveness and durability of protection against future SARS-CoV-2 infection conferred by COVID-19 vaccination and previous infection; findings from the UK SIREN prospective cohort study of healthcare workers March 2020 to September 2021"

Supplementary Figure iii: Number of primary infections (A) and reinfections (B) in the SIREN cohort and national UK cases (C) by vaccine status and variant-dominant period, 07 December 2020 to 21 September 2021

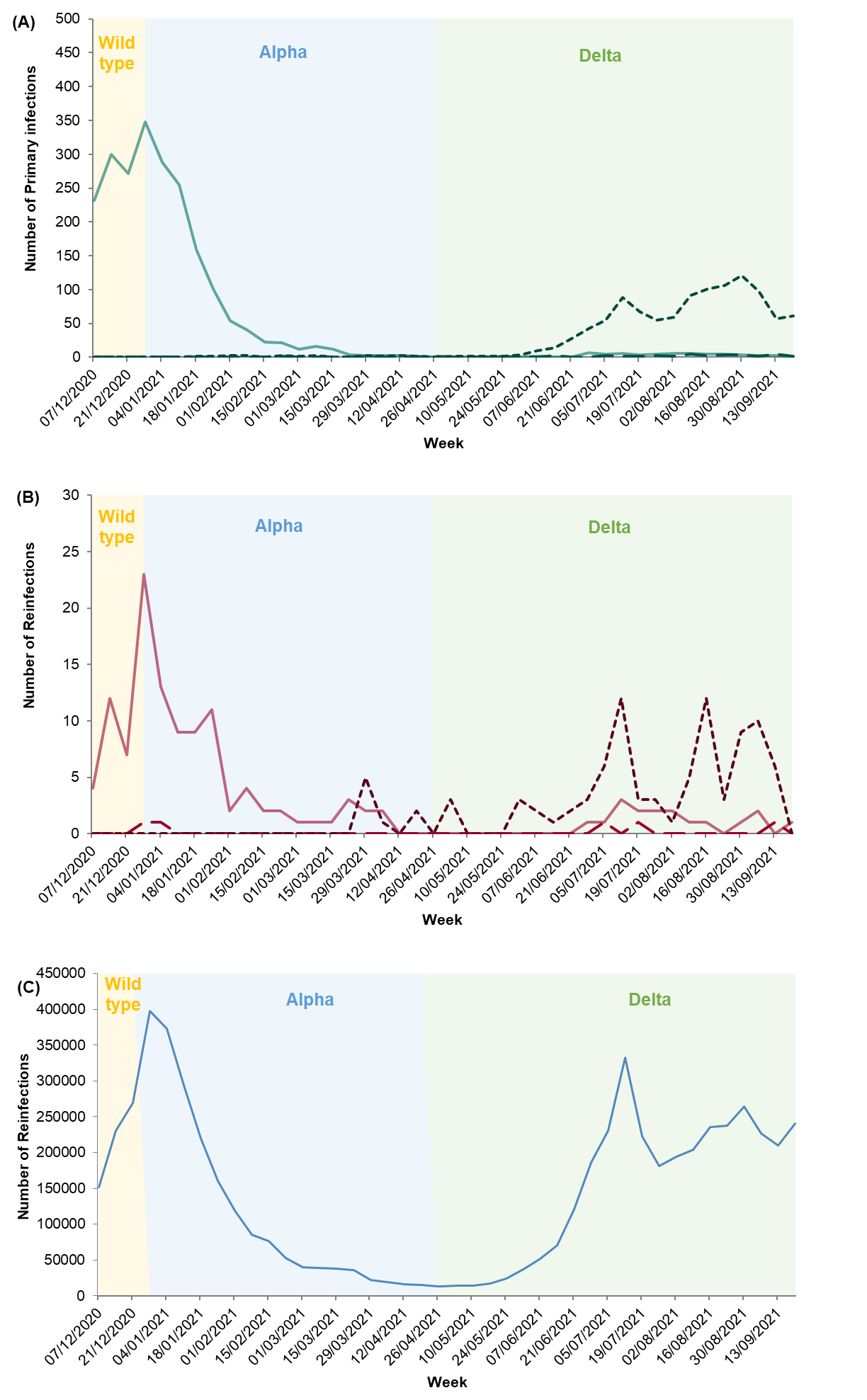

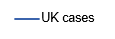

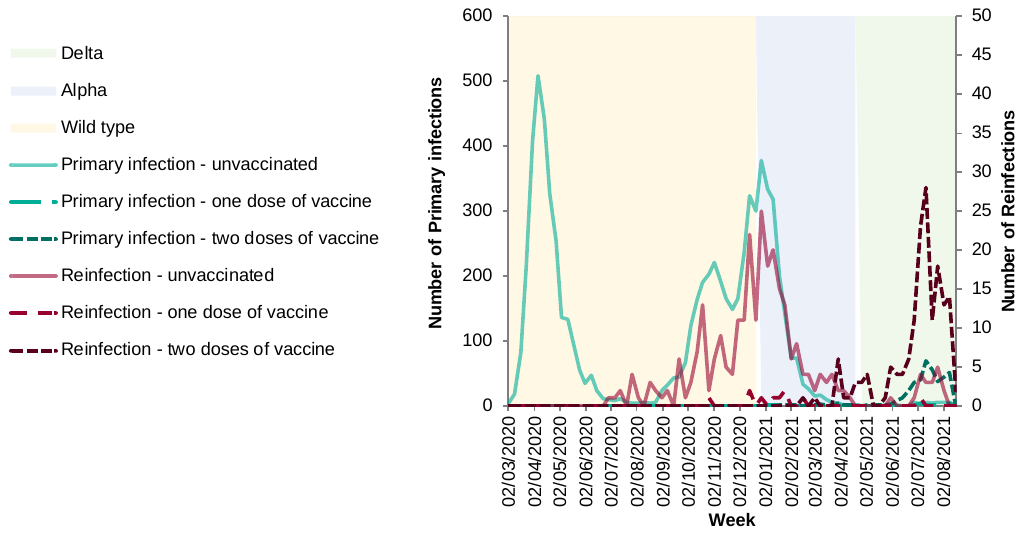

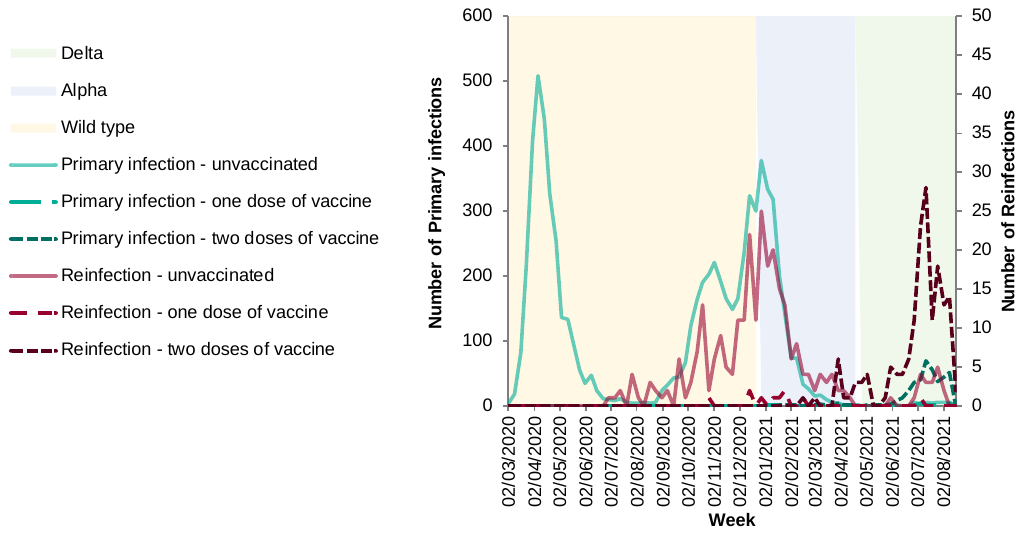

Data for panel C) Source: GOV.UK Coronavirus (COVID-19) in the UK
