## Supplementary Figure ii for "Effectiveness and durability of protection against future SARS-CoV-2 infection conferred by COVID-19 vaccination and previous infection; findings from the UK SIREN prospective cohort study of healthcare workers March 2020 to September 2021"

Supplementary Figure ii: Description of cohort enrolment, vaccination coverage and SARS-CoV-2 variant dominance during analysis period, June 2020 to September 2021

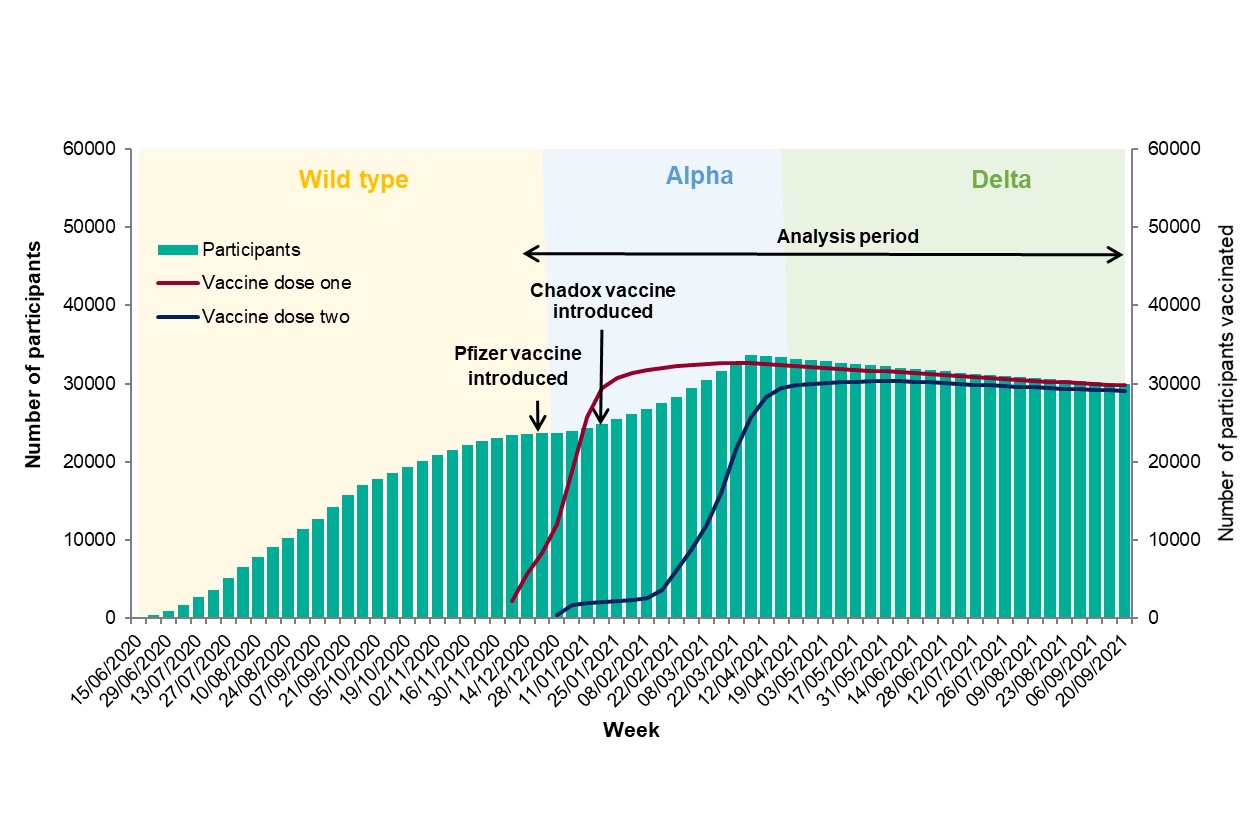
